# Examining the Health Impacts of Climate Change Through Electronic Health Records: A Rapid Review

**DOI:** 10.1101/2025.08.27.25334567

**Authors:** Arun Dahil, Cameron Pinn, Lucy Smith, Nehal Hassan, Nuno Koch Esteves, Glenn Simpson, Hajira Dambha-Miller

## Abstract

**Introduction:** Temperature extremes, including elevated heat and cold, are important environmental determinants of health whose frequency and duration are increasing due to climate change. Ecological and time-series studies have established links with adverse outcomes but often lack individual-level detail. Electronic health records (EHR) provide an alternative source, yet their use in climate-health research remains inconsistent.

**Methods:** We conducted a rapid review of peer-reviewed studies using EHR data to examine associations between temperature extremes and health outcomes across healthcare settings. The aim was to assess how health impacts of temperature extremes have been captured and coded within EHR-based research, and to identify methodological and coding-related gaps. Searches of seven databases identified eligible studies, and data were extracted on exposure definitions, outcome coding, methods, findings, and limitations.

**Results:** Of 1,616 records identified, 526 duplicates were removed, leaving 1,090 for screening; 58 studies met inclusion criteria. Extreme heat was most frequently studied, with fewer analyses of cold. Common outcomes included morbidity, cardiovascular admissions, asthma, and pregnancy-related conditions. Mental health outcomes were rarely examined, subgroup analyses were mostly age-based, and studies focused on high-income countries. Exposure metrics and coding practices varied widely, with limited reporting of diagnostic codes and individual-level mediators.

**Conclusion:** Harmonized exposure definitions, broader outcome coverage, and integration of socio-demographic and individual-level factors are needed to strengthen EHR-based climate-health research and guide targeted interventions.

## Introduction

Temperature extremes, including prolonged periods of high heat or cold, are increasingly recognized as important environmental determinants of health. Their frequency and duration are projected to rise due to climate change, with significant implications for healthcare demand and health outcomes across diverse population groups [1]. Numerous studies have documented associations between extreme temperatures and a range of adverse outcomes, including increases in emergency department visits, hospitalizations, and mortality [2,3]. These effects are disproportionately observed in older adults, individuals with chronic health conditions, and residents of socioeconomically disadvantaged areas [4–6]. Despite these findings, most existing research relies on ecological or time-series designs using aggregated health data. While these approaches have advanced understanding of short-term effects, they are limited in their ability to capture individual-level variation, account for pre-existing health conditions, or consider how patterns of healthcare utilization and behavioral responses may mediate climate-health relationships. This restricts the capacity to identify the most vulnerable subpopulations and develop targeted, evidence-based interventions.

Electronic health records (EHR) offer an alternative and potentially more powerful approach. These routinely collected datasets provide longitudinal, individual-level information on diagnoses, prescriptions, consultations, and demographic characteristics. Large-scale resources such as the Clinical Practice Research Datalink (CPRD) and the Secure Anonymised Information Linkage (SAIL) Databank hold millions of records, enabling population-based analyses of climate-related health outcomes [7]. However, EHR-based studies in this field are hampered by substantial methodological heterogeneity. Differences in temperature metrics, exposure windows, and diagnostic coding systems, combined with inconsistent reporting of these methods, limit interpretability, comparability, and synthesis of findings. Furthermore, research to date has tended to focus on a narrow range of health outcomes and high-income settings, often neglecting mental health and other potentially climate-sensitive conditions. Accordingly, this rapid review aims to examine how health impacts of temperature extremes have been captured and coded within EHR-based research across healthcare settings, and to identify methodological and coding-related gaps in the current evidence base.

## Methods

This rapid scoping review followed the methodological framework of Arksey and O’Malley [8], refined by Levac et al. [9], and adhered to the Preferred Reporting Items for Systematic Reviews and Meta-Analyses extension for Scoping Reviews (PRISMA-ScR) [10]. The process involved: (i) identifying eligibility criteria and relevant studies; (ii) selecting studies for inclusion; (iii) extracting and charting data; and (iv) collating, summarising, and reporting findings.

### Eligibility Criteria

We included studies that were: (i) based on EHR or linked routinely collected health data; (ii) examined health outcomes, healthcare utilization, or interventions in the context of temperature extremes (heatwaves, cold events, extreme humidity, or other temperature-related exposures); (iii) published in peer-reviewed journals, academic books, conference proceedings, or relevant grey literature; and (iv) available in English. Studies that did not meet these criteria were excluded.

### Search Strategy

Searches were developed in consultation with an information specialist from the University of Southampton and refined with input from climate change and medical experts. Initial terms were piloted and revised before final implementation. Keywords and Medical Subject Headings (MeSH) combined terms for exposures (e.g., “temperature extreme*,” “heatwave*,” “hot weather,” “cold spell*,” “freeze*,” “extreme humidity,” “climate”) with health data sources (e.g., “EHR,” “routinely collected data,” “registry data,” “health administrative data”). Variants were searched using title, abstract, and indexing fields. Searches were run from database inception to 7 July 2025 in MEDLINE (Ovid), Embase (Ovid), GreenFILE, Web of Science, Scopus, CINAHL (EBSCOhost), and Cochrane.

### Study Selection

Results were imported into Rayyan and duplicates removed. Titles and abstracts were screened independently by three reviewers (CP, AD, LS). Full-text screening was conducted for studies meeting inclusion criteria or where eligibility was unclear, with disagreements resolved by consensus or adjudication by a third reviewer.

### Data Extraction and Charting

A standardized, piloted data extraction form was used to record: study characteristics (authors, year, country, design); population demographics; exposure type, metrics, and duration; health outcomes (morbidity, mortality, hospital admissions, primary care visits, or other utilization measures); EHR dataset and meteorological data source; coding approach (diagnostic, symptom, or utilization codes, classification system used); specific codes; key findings; and reported limitations. Data were extracted by CP, AD, LS, NH, and NKE.

### Data Synthesis

Extracted data were summarized descriptively and grouped thematically by type of climate variable studied, health outcomes assessed, exposure definitions and metrics, codes and terminology used, sources of climatic data, and patterns in study design and population characteristics. Key findings and limitations were analysed to identify trends, methodological issues, and evidence gaps, summarized narratively and presented in summary tables.

## Results

### Study Selection

Database searches yielded 1,616 records; 526 duplicates were removed, leaving 1,090 for title and abstract screening. A further 845 were excluded, and 245 underwent full-text review. One hundred and eighty-seven were excluded, resulting in 58 studies for analysis (Figure 1).

**Figure 1:**
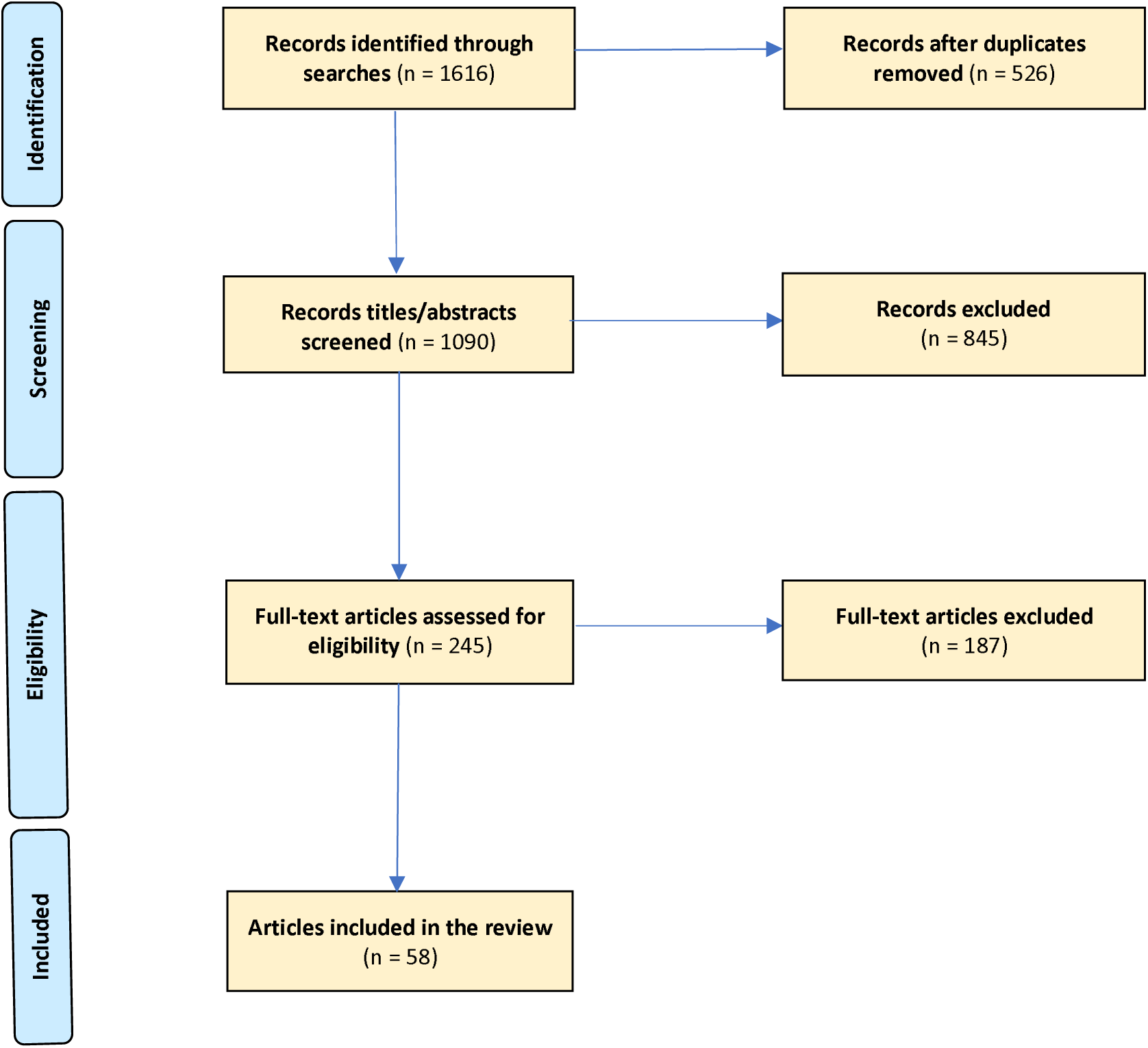
Adapted PRISMA Flow Chart showing explaining the study’s documentary inclusion process (Examining the Health Impacts of Climate Change Through Electronic Health Records: A Rapid Review)

### Study characteristics

Full details on all study characteristics can be found in supplementary data, table 1. Of the 58 studies, 42 (72%) were from high income countries [11,13,14,15,16,18,22,23,24,25,26,27,29,30,31,32,33,36,37,39,40,41,42,44,45,46,47,49,50,51,52,53,54,55,56,57,59,61,63,64,65,66], 11 (19%) were from upper middle-income countries [17,19,20,21,28,35,43,48,58,62,68] and three (5%) were from lower middle-income countries [12,34,67]. Two studies (4%) were extracted from a mix of low and high income countries [38,60].

In total, 21 studies were located in the USA [13,14,15,16,18,22,25,26,29,30,31,33,39,42,47,50,52,53,63,64,65], 10 focused on China [17,19,20,21,28,35,43,48,58,68], four in the UK [32,51,54,59], three in Canada [11,44,57], two in Spain [27,40], Australia [45,55], Italy [36,49], Finland [24,46], and one study each from Ethiopia [67], Macao territory [66], India [62] France [61], Belgium [23], Kuwait [56], North Tanzania [12], Singapore [41], Vietnam [34] and Japan [37]. Lastly, two studies used mixed country populations [38,60]. Figure 2 visually displays the countries of origin.

**Figure 2:**
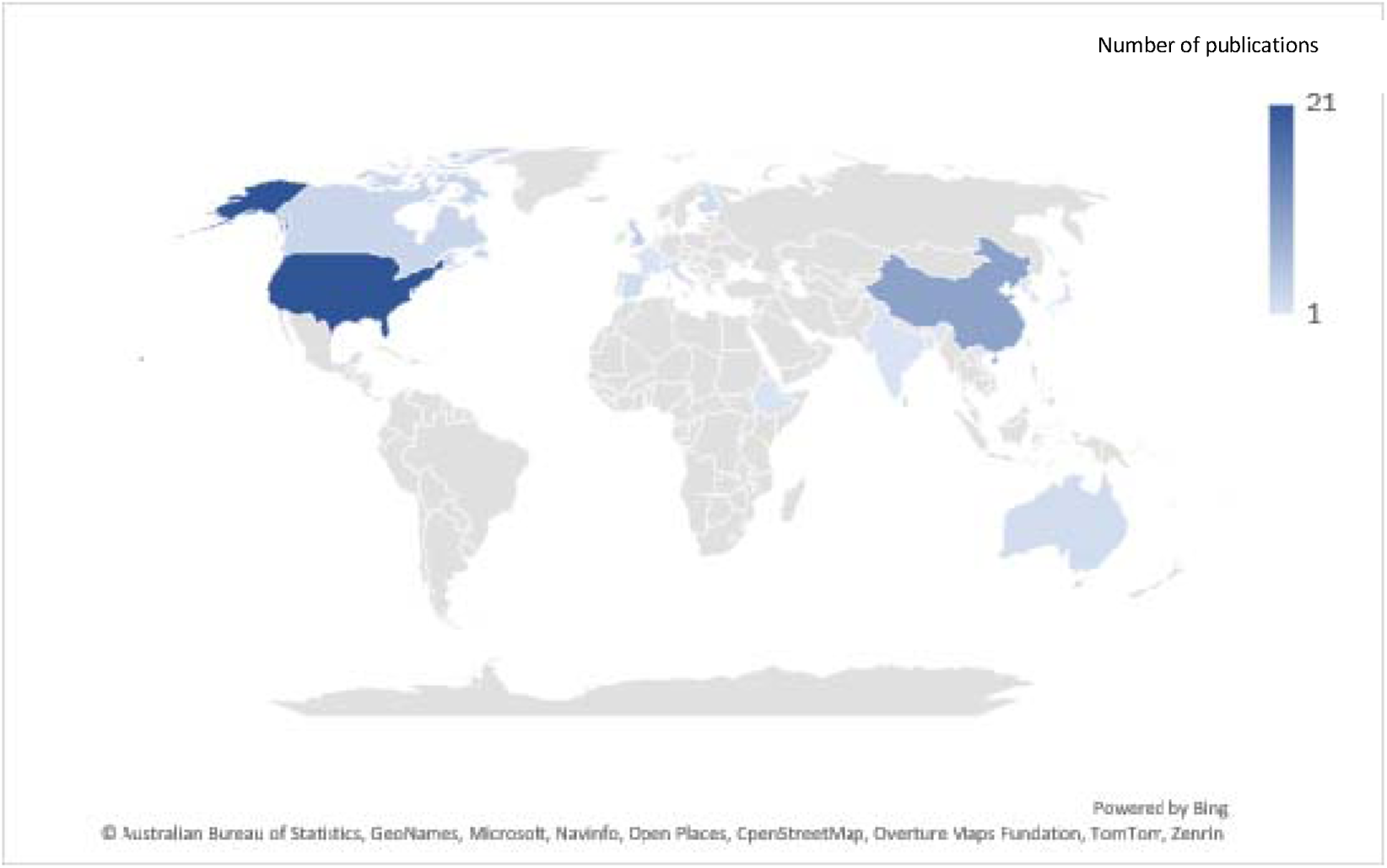
A heat map of geographic location for all studies included in the rapid review

There has been an increasing trend in publications post 2020, with 45 papers (77%) being published between 2020-2025, as shown in figure 3. The main study type used was a retrospective cohort design (n=33) and the look back period ranged from two to 24 years, with a mean of 9.5 years.

**Figure 3:**
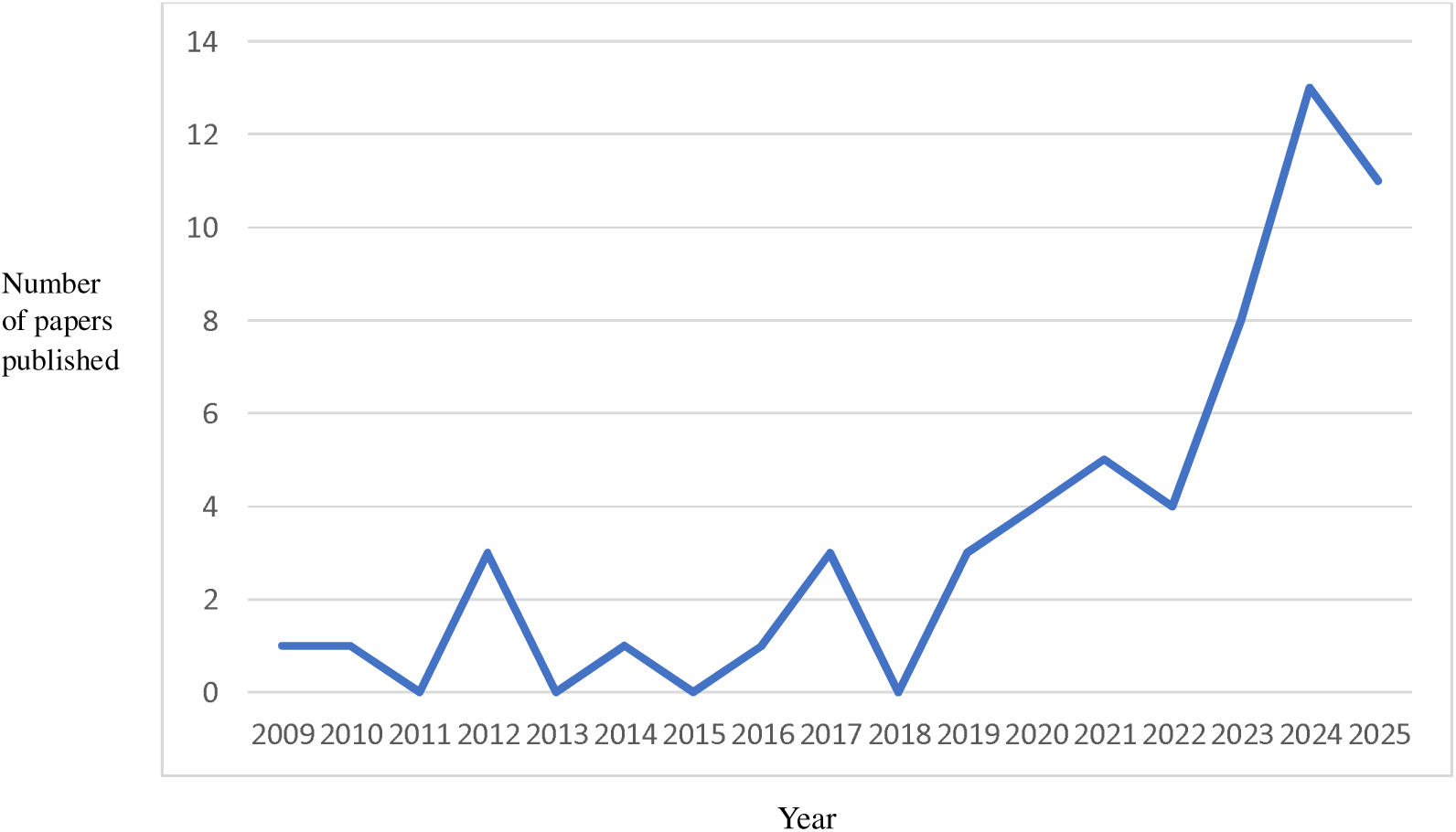
A line frequency graph showing number of publications by year which are focused on climate change exposures and health outcomes variables

Four studies made predictions based on forecast models [13,14,33,37], one study included a climate change intervention [44] and one study was a systematic review [60].

Populations included in the studies were mostly adult, although some studies included on children (n=10). No studies referred to subgroup analysis beyond age as a factor. In terms of other sub-populations, the impact of climate extremes on veterans were also analysed (n=2). Many studies referred to whole cohorts, and the demographic details of who was in this cohort were not made explicit.

### The use of electronic health care records to record health outcomes

EHR were used to capture health outcomes across a broad range of themes, including mortality (n=13), neonatal/pregnancy outcomes (n=13), health service usage/cost (n=11), reference to a specific illness or disease area/s (n=23), temperature related illness/injuries (n=5), mental health (n=4), (two of which referred to post-partum depression) outcomes after surgery (n=1). For studies which made reference to a specific illness or disease area, the majority of studies focused on cardiovascular disease (n=8), asthma (n=5), respiratory diseases (n=4), temperature related illness or injuries such as dehydration, heat stroke or frost bite (n=6), mental health (n=4), ischemic stroke (n=2), gestational diabetes (n=2) acute kidney disease (n=1), infectious diseases (n=1). Health outcomes related to neonatal/pregnancy outcomes, included still births (n=2), placental abruption (n=1), growth and/or birth defects (n=3), placental abruption (n=1), maternal morbidity (n=1), and premature delivery (n=1). No study considered the impact of climate change exposure on people with multiple long-term conditions. Figure 4 provides a visual representation of the different health outcomes explored using EHR.

**Figure 4.**
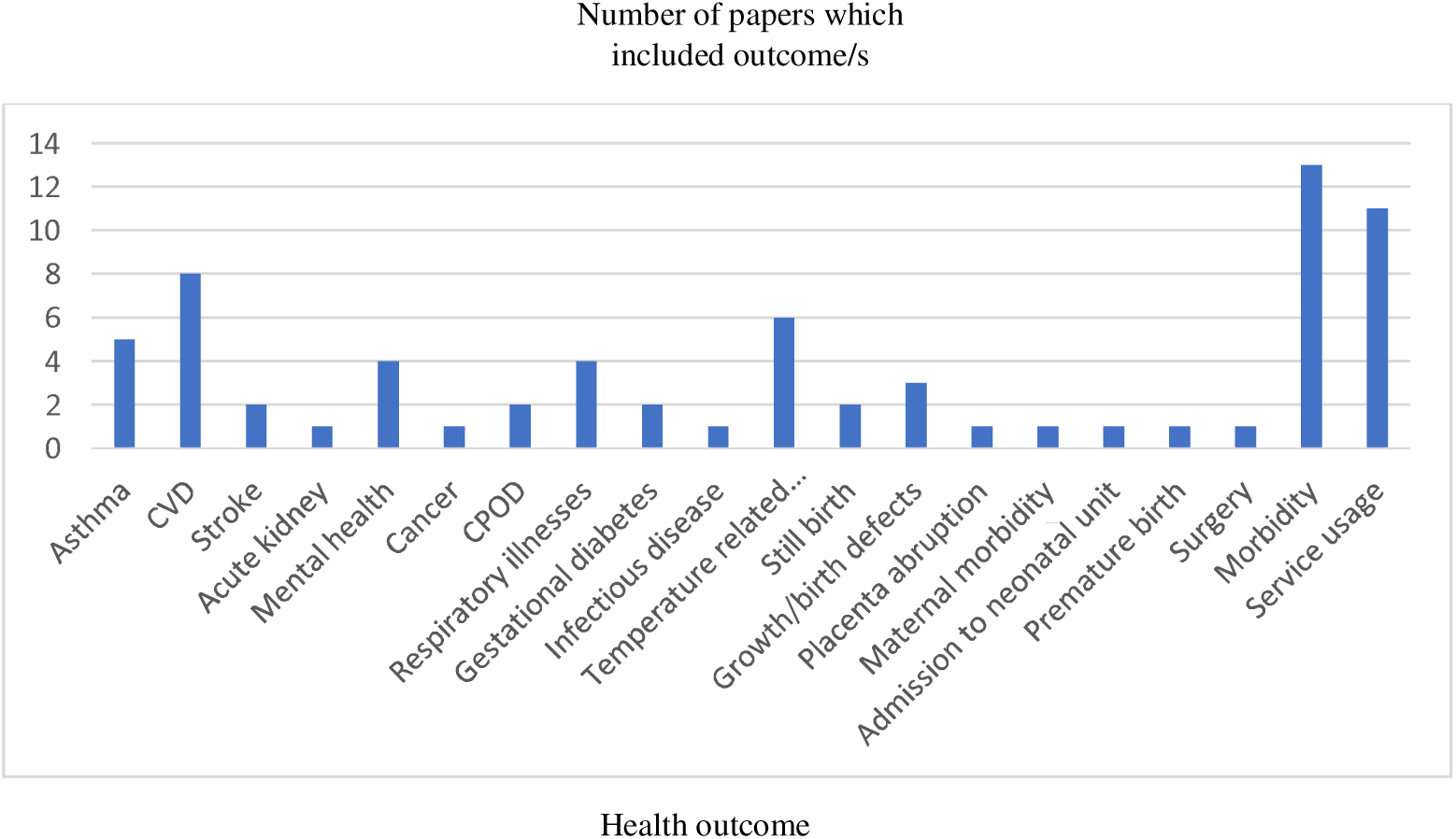
A bar graph to show health outcomes extracted from the EHR for all studies focused on climate change exposure variables and health outcomes

The majority of studies used either ICD 9/10 codes to extract the data from EHR (n=31), 16 studies used different registries to obtain their data, two studies used SNOWMED codes, and nine studies did not make explicit how they extracted their data. There was a general missingness of coding detail across the studies, and in particular where ICD 9/10 or SNOWMED were used, the specific codes utilized to extract the data were only included in 15 papers.

Lastly, the settings from which EHR data was extracted was primarily secondary hospital level data, especially from emergency departments. Only one study examined primary care data, and this was in relation to missed appointments [65], and one study looked at visits to a tertiary mental health clinic [57].

### Climate change variables

Twelve different exposure variables were extracted from the 58 papers, these included microclimate data, thunderstorms/major storms, flooding, snowfall, solar output, wind speed, humidity, rainfall/precipitation, air pollutants, extreme cold weather, extreme hot weather, and temperature including both extreme heat and cold weather events. The most common exposure variable, featured in 29 (50%) of the papers was temperature, followed by a focus on extreme heat events (n=13). Cold weather events were less frequently focused on (n=2). Table 1 shows the number of papers for each climate change variable.

**Table 1:** Number of papers examining each climate change exposure variable.

| **Climate change exposure variable** | **Number of papers** | **Paper reference** |
| --- | --- | --- |
| Microclimate | 2 | 26, 31 |
| Storms/thunderstorms | 2 | 28, 29 |
| Flooding | 2 | 24, 29 |
| Solar output | 2 | 53, 66 |
| Snowfall | 1 | 64 |
| Wind speed | 2 | 49, 66 |
| Humidity | 8 | 29, 33, 34, 49, 52, 53, 56, 66 |
| Precipitation | 6 | 35, 37, 42, 49, 53, 65 |
| Air pollutants | 8 | 15, 19, 20, 21, 22, 61, 66, 73 |
| Extreme cold weather events | 2 | 27, 50 |
| Extreme heat weather events | 13 | 10, 13, 15, 18, 23, 24, 31, 36, 41, 46, 56, 61, 62 |
| Temperature (both hot and cold) | 29 | 2, 12, 14, 16, 17, 19, 20, 25, 26, 32, 33, 34, 35, 37, 38, 39, 40, 43, 45, 48, 51, 52, 54, 57, 58, 59, 60, 63, 65 |

Definition and metrics used across exposure variables varied across papers. For definitions of extreme heat, studies used different percentile cut off points, ranging from a temperature over the 90^th^ percentile, to over the 99^th^ percentile, though the 97.5^th^ percentile was most frequently cited as a suitable cut off point for an extreme heat threshold (n=4). Studies also varied in the number of days a certain temperature had to be experienced for it to be classified as a heat wave (between 2-4 days). Similarly, variance was seen in cut-off points for extreme cold, with temperatures ranging from below the 2.5^th^ percentile, to below the 10^th^ percentile. Inconsistency in duration of temperature recordings was seen, with recordings ranging from hourly (n=4), daily (n=17) weekly (n=3), monthly (n=2) to annually (n=2). Time lag data from exposure to health outcome also differed, from 24 hours, 14 days, 21 days, 30 days to 30 years. Lastly, four papers [22,29,61,68] referred to using a resolution grid to provide a spatial imagery of temperature, and areas selected varied from 1×1 km to 500 km.

### Key findings by exposure variables and health outcomes

Table 2 provides a summary of key findings for each study. Overall, the findings from the narrative synthesis indicate that extreme temperatures, either cold or hot, can impact health across a range of health outcomes, and this can vary by gender and deprivation, with those who are most socially vulnerable experiencing increased risks.

**Table 2:**
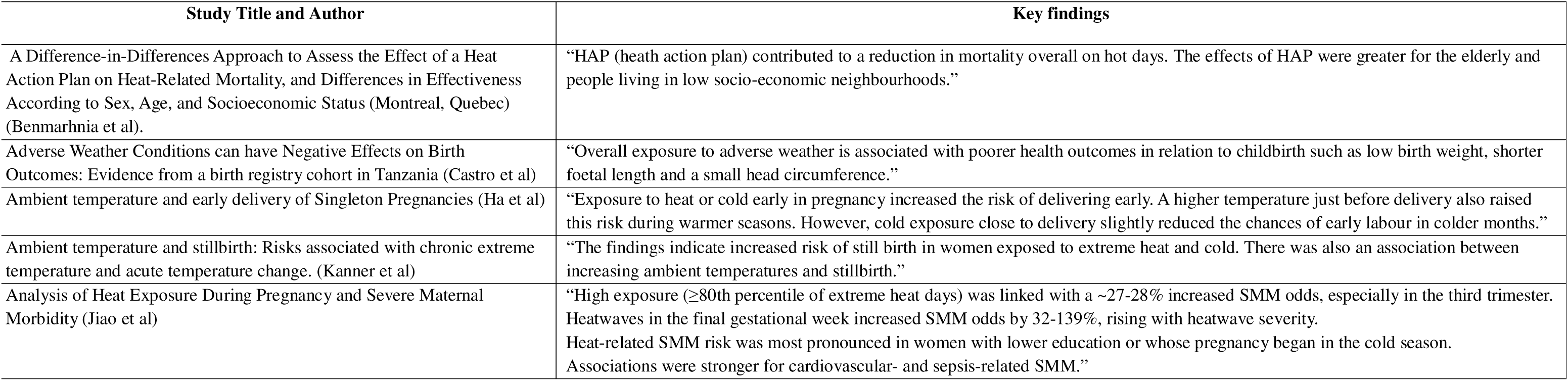
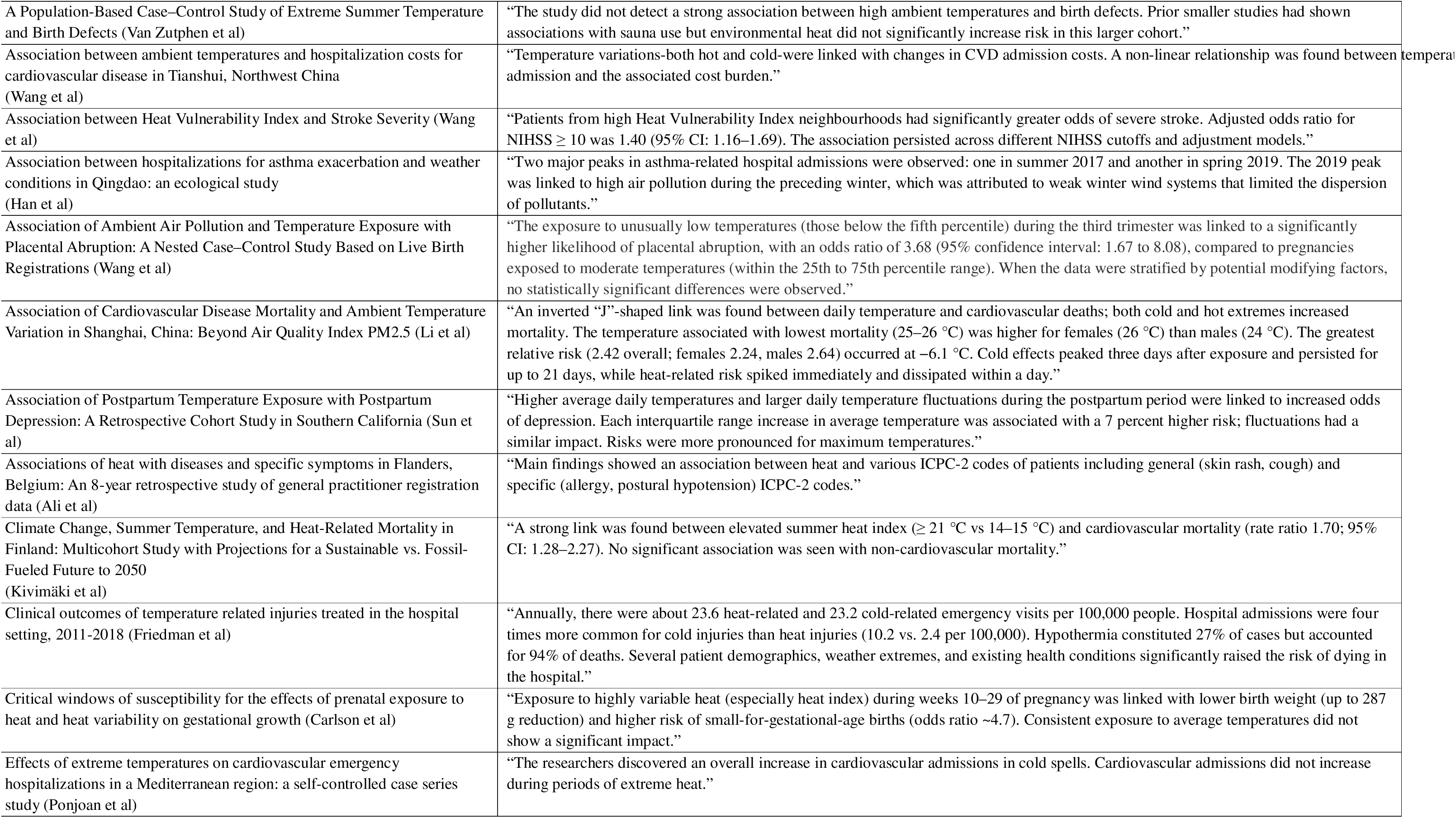
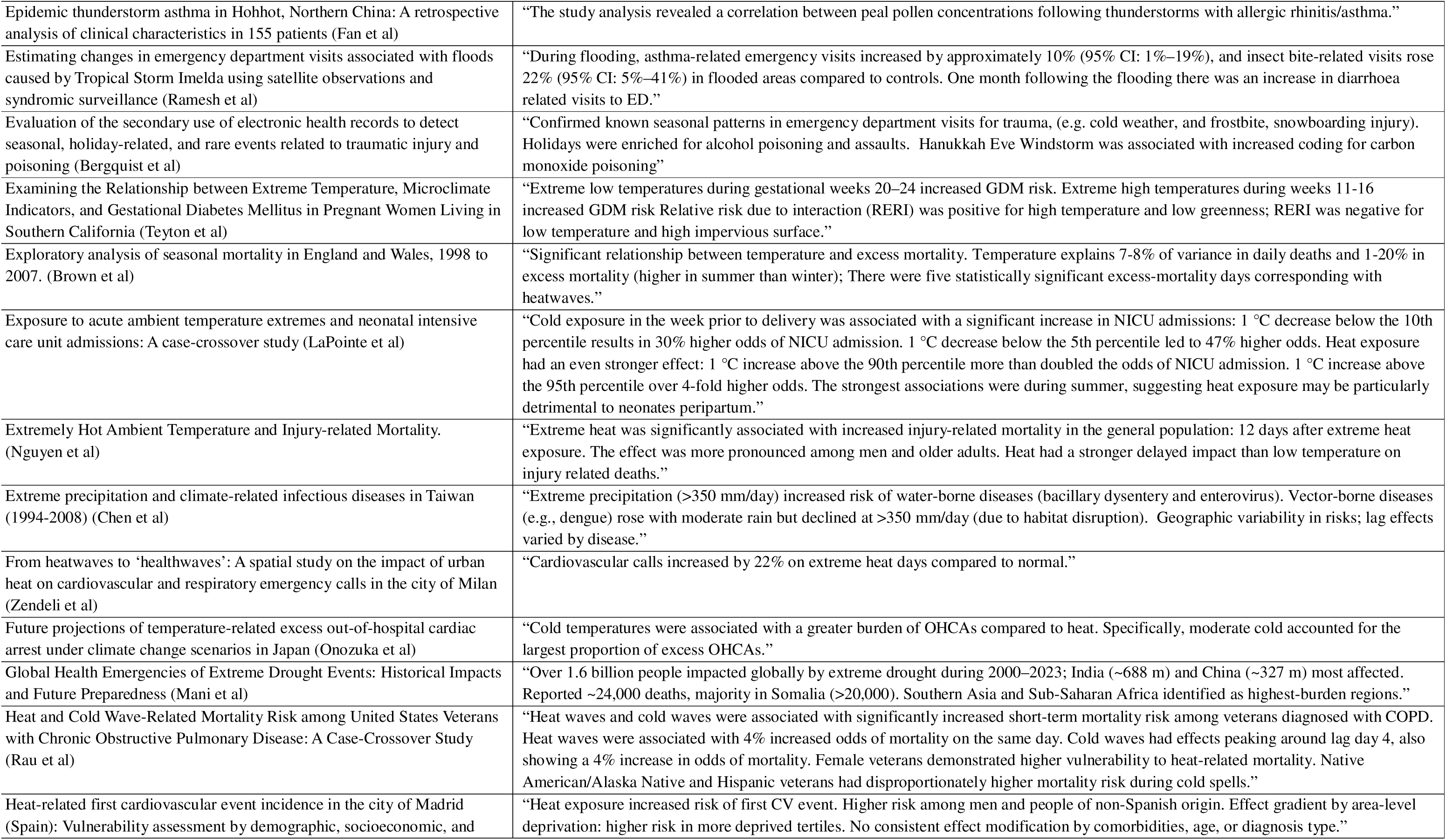
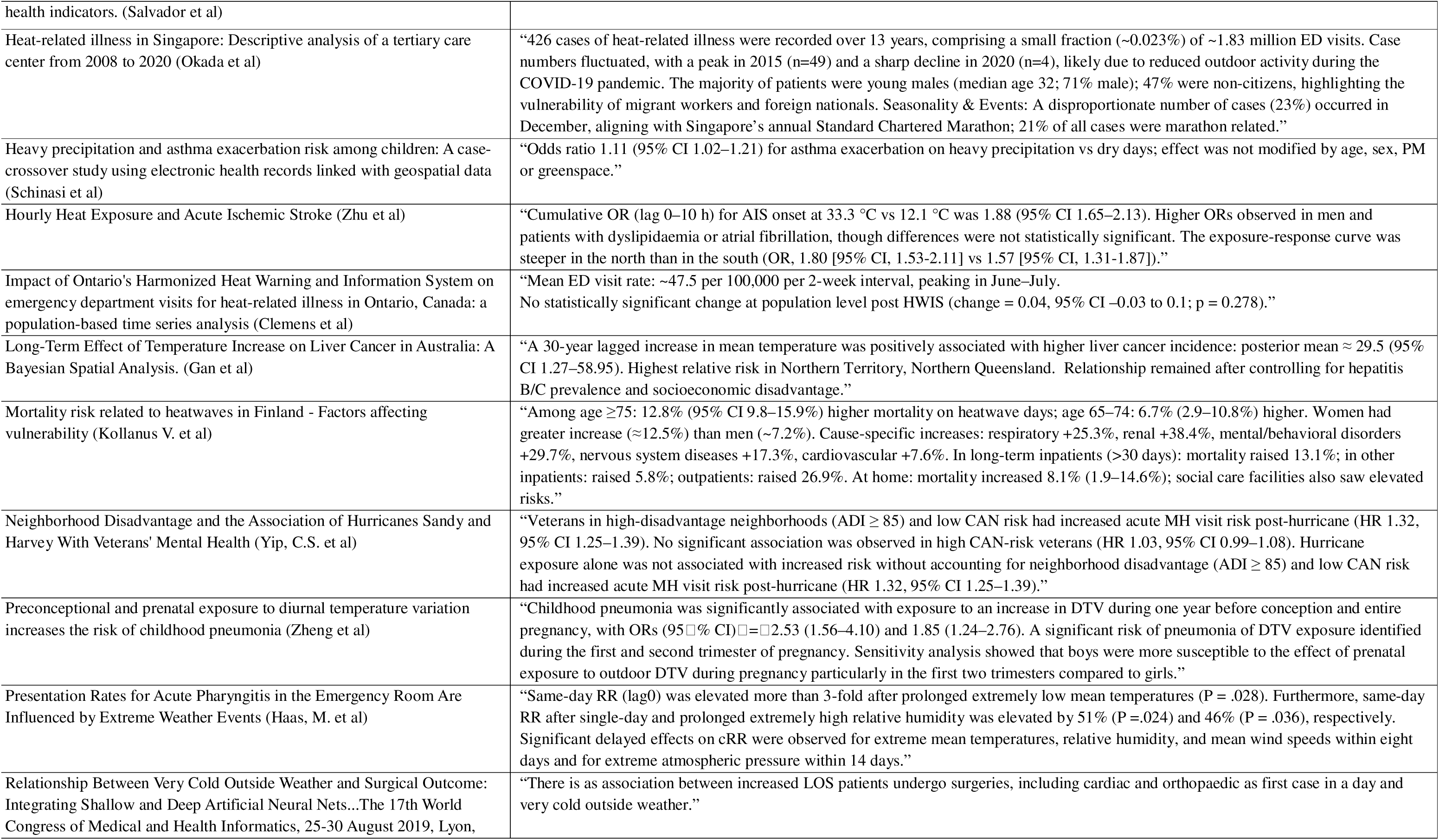
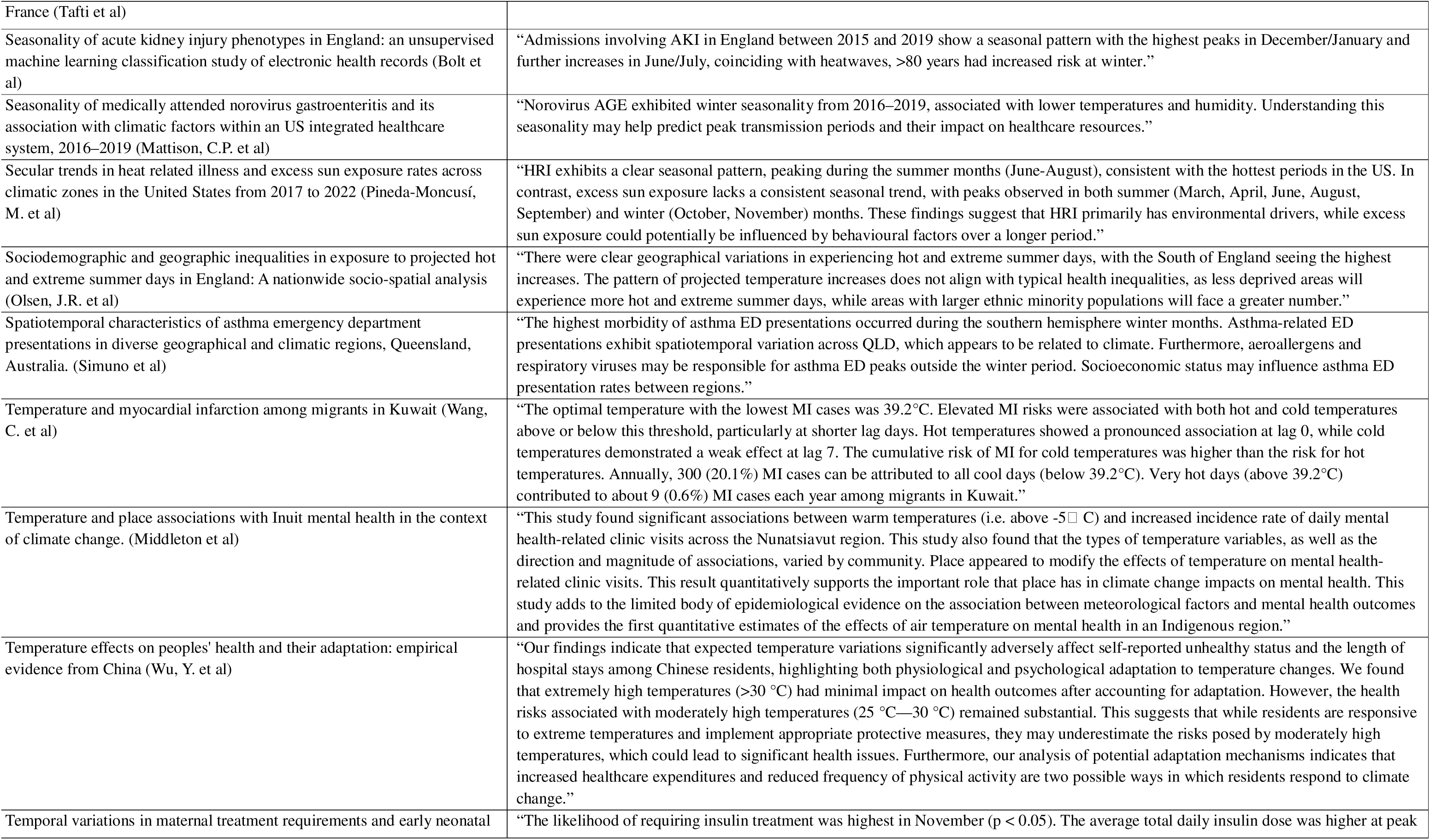
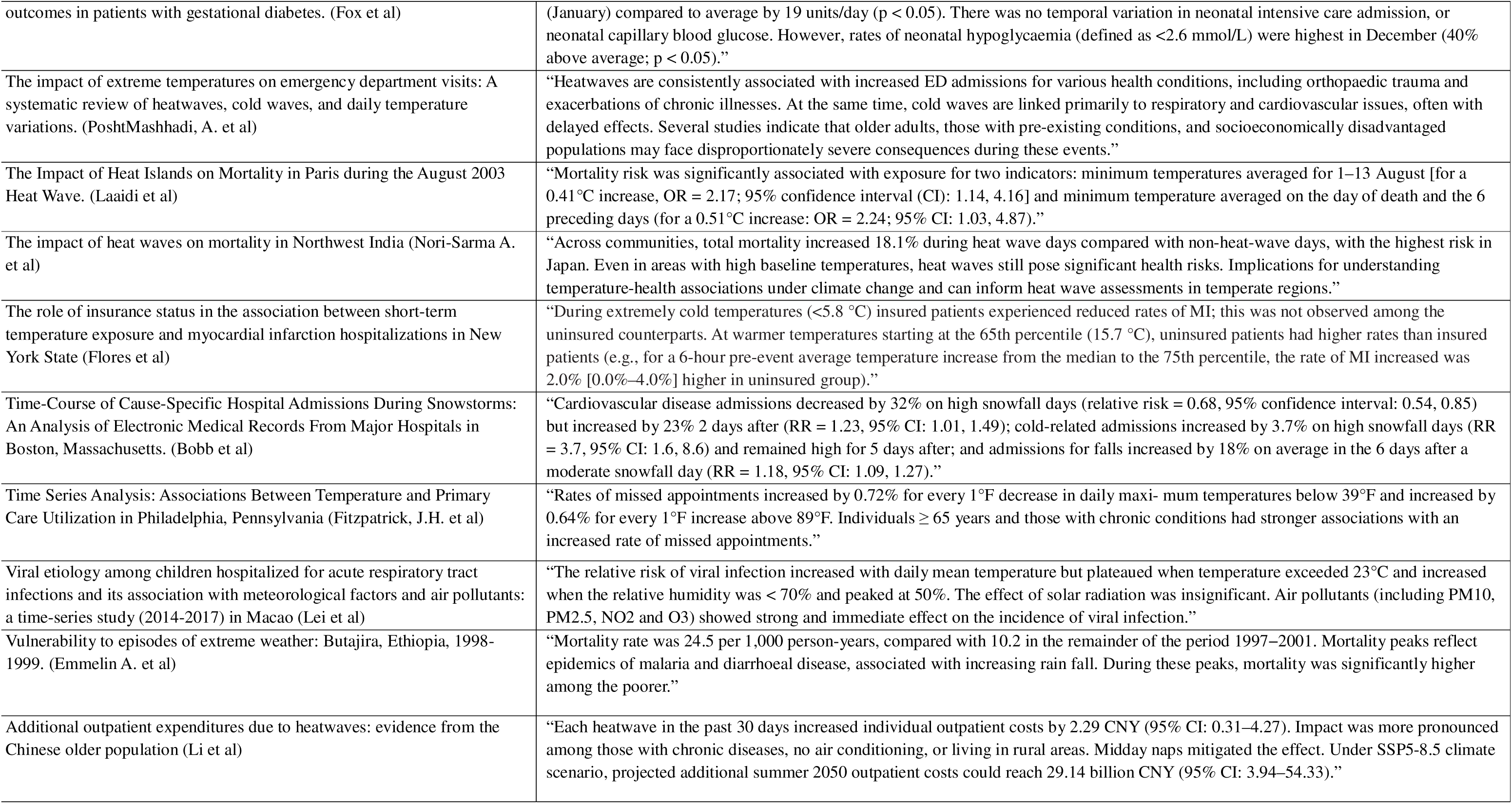
Key findings from extracted papers on the impacts of climate change exposure on health outcomes of patients.

In relation to neonatal outcomes, exposure to extreme temperature leads to poorer outcomes for baby, but these vary by hot and cold. Increased exposure to heat is associated with high risk of still birth, maternal morbidity, gestational diabetes mellitus and lower neonatal growth. Exposure to extreme cold was linked with placenta abruption and admission to the neonatal intensive care unit.

Across the papers focused on cardiovascular disease, an inconsistent pattern emerged, with some studies finding extreme heat increases CVD mortality and admissions, whilst others found that risk increased with extreme cold temperatures. Gender differences and deprivation also influenced this relationship. A more consistent pattern emerged for mental health service usage, which increased during hot temperatures.

Lastly, cold weather extremes were more likely to lead to hospital admissions for conditions such as hypothermia. An increase in mortality was consistent with extreme heat, with females and those from areas of deprivation being more at risk. Young males more at risk of heat-related admissions to hospital, which is likely linked to behavioral factors.

### Limitations in climate change exposure and health outcome research

A consistent set of limitations emerged across studies. Most excluded individual-level factors, such as uncoded chronic conditions, socio-demographics, and behavioral adaptations (e.g., using air conditioning during heat events). Missing data were common due to retrospective designs, limiting follow-up and potentially omitting cases not presenting to emergency departments. Geographic coverage was often narrow, with many single-city or urban-focused studies. Accuracy of environmental exposure data varied by country, and climate models relied on assumptions that may not hold. Finally, ecological designs limited causal inference.

## Discussion

In this study, we aimed to identify gaps in how the health impacts of climate extremes are captured and coded in EHR-based research. We found that while publications have grown over the past five years, most originate from the USA and China, with none from low-income countries, despite higher vulnerability. Research is largely based on retrospective hospital EHR, heavily reliant on ICD-9/10 codes without consistent specification, and focused on extreme heat, with little on extreme cold and inconsistent exposure definitions. Health outcomes are predominantly cardiovascular, respiratory, neonatal, and pregnancy-related, with limited attention to mental health, multimorbidity, and primary or community care. Few studies include individual-level risk factors or social vulnerability analyses. Future research should expand geographic scope, harmonize definitions, improve transparency, and include more diverse settings, disease areas, and high-risk populations.

### Comparison with existing literature

This review confirms the limited attention given to mental health outcomes in relation to climate extreme exposures, consistent with previous findings [69]. Given that those most vulnerable to climate extremes are also at higher risk of mental health issues, this represents an important evidence gap [70,71]. Our results also align with literature showing that children, particularly newborns and infants, face elevated risks from climate change [72,73]. However, we identified only 10 studies on this group, primarily addressing neonatal or respiratory outcomes; further research should examine climate extremes across the full developmental trajectory [74].

A notable finding was the lack of methodological consistency in EHR-based climate-health research, also observed in other domains such as mental health [75,76]. Most studies applied retrospective time-series cohort designs, which are common in environmental health but limited in their ability to account for unmeasured confounders, including individual-level mediators of exposure effects [76]. Heterogeneity in climate exposure measures was rarely addressed, and few studies incorporated social vulnerability indicators such as education, housing, income, location, or access to care [77]. Similarly, although people with chronic conditions are more susceptible to climate extremes [78], none of the included studies examined the implications of multimorbidity, likely underestimating health impacts in these high-risk populations.

### Strengths and limitations

This rapid review provides a structured synthesis of how temperature-related health impacts have been examined using electronic health records, offering a timely overview of current practices in exposure definition, outcome coding, and data utilization. Strengths include a comprehensive, multi-database search strategy developed with an information specialist, ensuring broad coverage of peer-reviewed literature, and adherence to established scoping review frameworks and PRISMA-ScR reporting standards, enhancing transparency and reproducibility.

Limitations include restriction to English-language publications, potentially excluding relevant research from non-English speaking regions; omission of grey literature; and the inherent constraints of a rapid review, such as the absence of full quality appraisal and exclusion of evidence published after the search cut-off date. In addition, the focus on temperature-related extremes means other climate hazards (e.g., flooding, wildfires, droughts) were not captured, limiting the broader applicability of findings across the full spectrum of climate–health interactions.

## Conclusion

Our review findings suggest that EHR-based climate-health research is expanding but remains geographically narrow, methodologically inconsistent, and limited in its inclusion of high-risk populations. Future studies should extend to low-income and climate-vulnerable regions; incorporate primary care, community, and mental health settings; and address neglected areas such as mental health, paediatric life-course outcomes, and multimorbidity. Harmonized exposure definitions and fully specified code lists are essential for comparability, while prospective designs linking EHR with environmental and socio-demographic data, and integrating individual-level mediators and social vulnerability metrics, will generate evidence that can inform targeted, equitable clinical and public-health interventions. Addressing these gaps is critical to guide effective policy and strengthen health system resilience in the face of intensifying climate extremes.

## Data Availability

All data produced in the present study are available upon reasonable request to the authors

## Ethics approval and consent to participate

Ethical approval was not required for this scoping review and therefore not applicable.

## Availability of data and materials

Data used during the current study are available from the corresponding author on reasonable request.

## Author Contribution Statement

AD: Writing - original draft; Investigation; Formal analysis. CP: Writing - original draft; Investigation; Formal analysis. LS: Writing - original draft; Supervision; Investigation; Formal analysis. NH: Investigation; Formal analysis. NKE: Investigation; Formal analysis. GS: Formal analysis; Writing - review and editing. HDM: Conceptualization; Writing - original draft; Writing - review and editing.

## Guarantor

HDM is guarantor.

## Acknowledgements

None.

Author’s information: None.

## Consent to publish

Not applicable.

## Declaration

The lead author affirms that the manuscript is an honest, accurate, and transparent account of the study being reported; that no important aspects of the study have been omitted. The opinions, results, and conclusions reported in this article are those of the authors and are independent from the funding sources.

## Appendix

### Supplementary material

**Supplementary Table 1:**
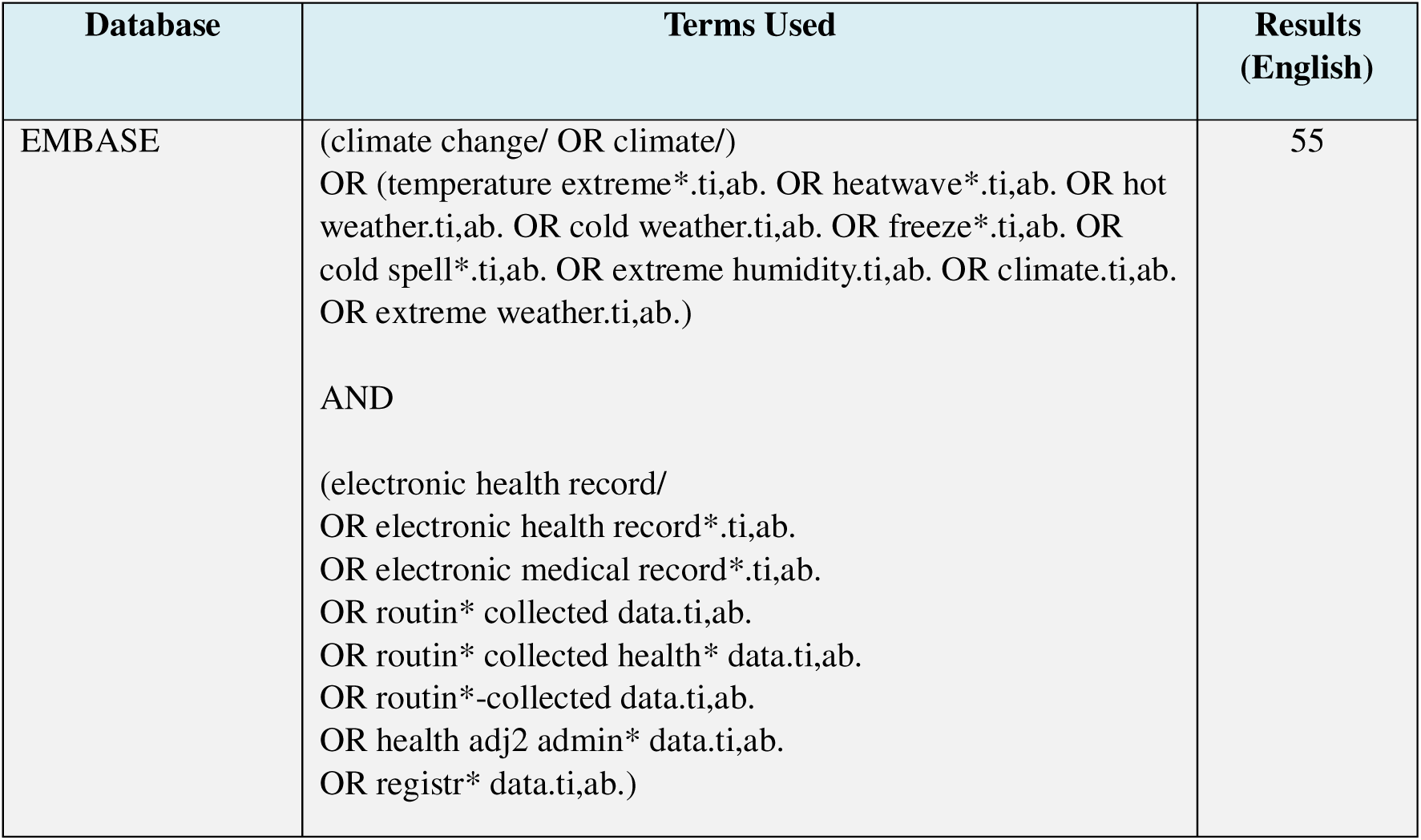
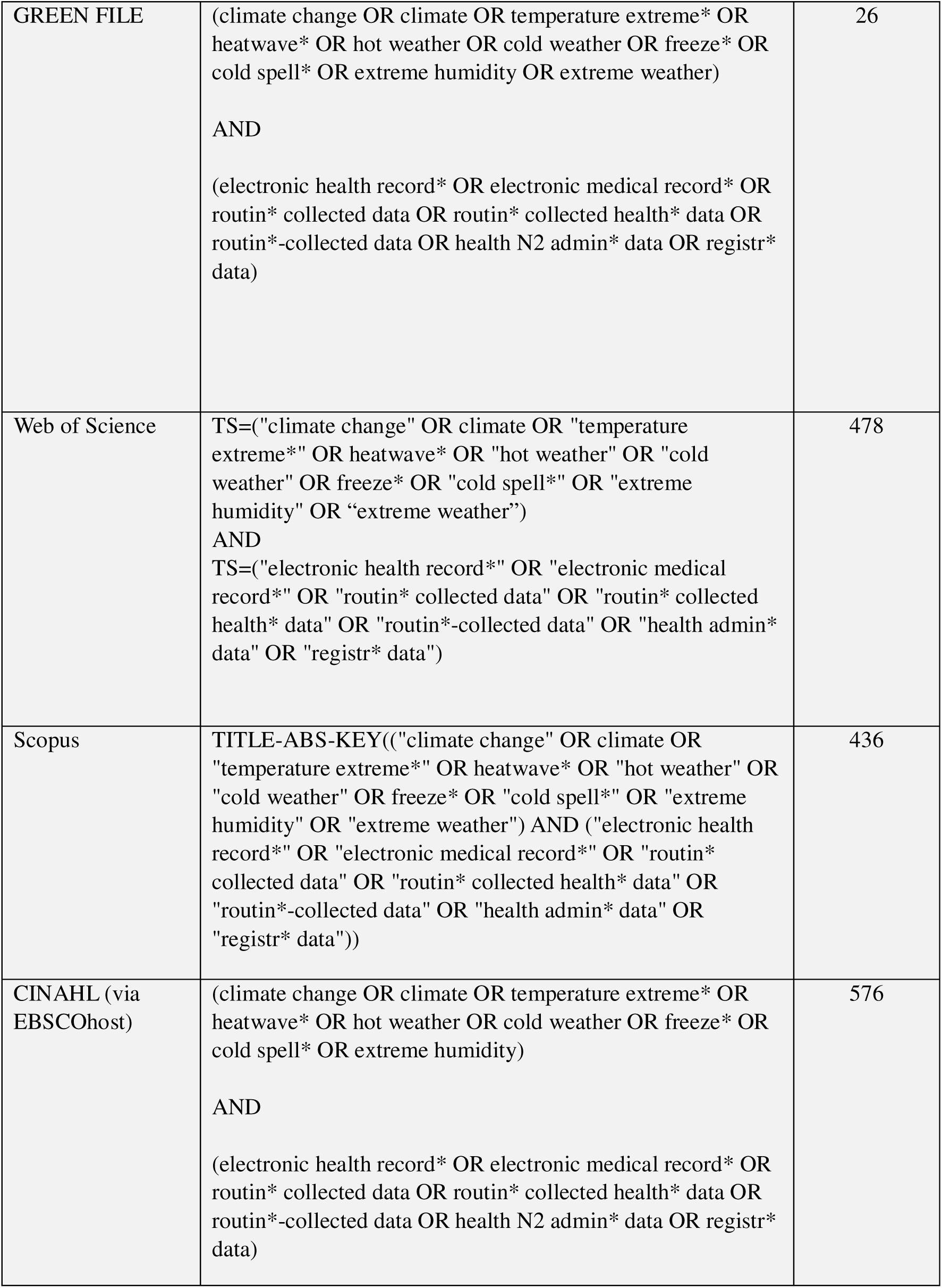
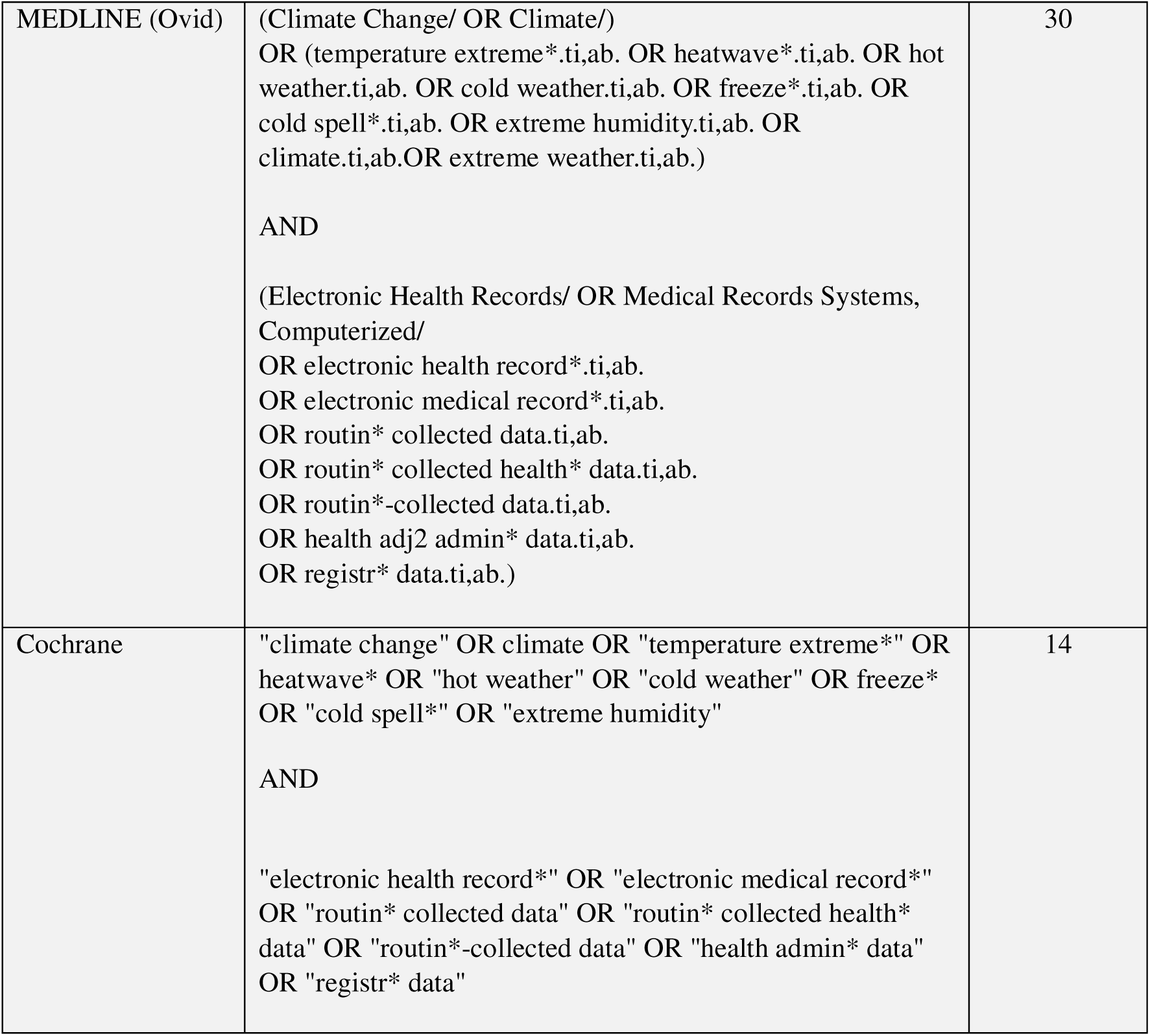
Search terms and number of papers extracted by databas.

**Supplementary Table 2:**
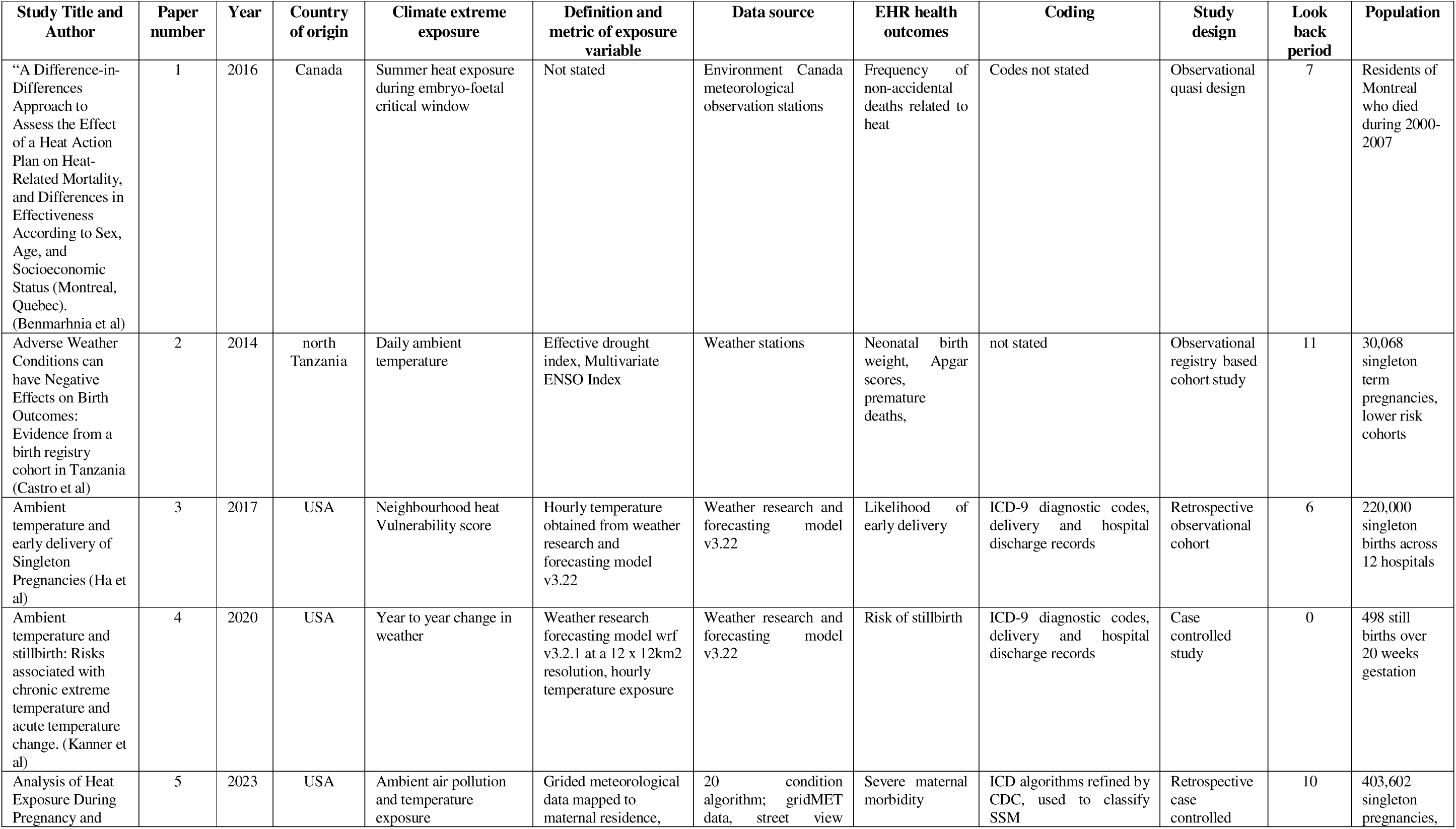
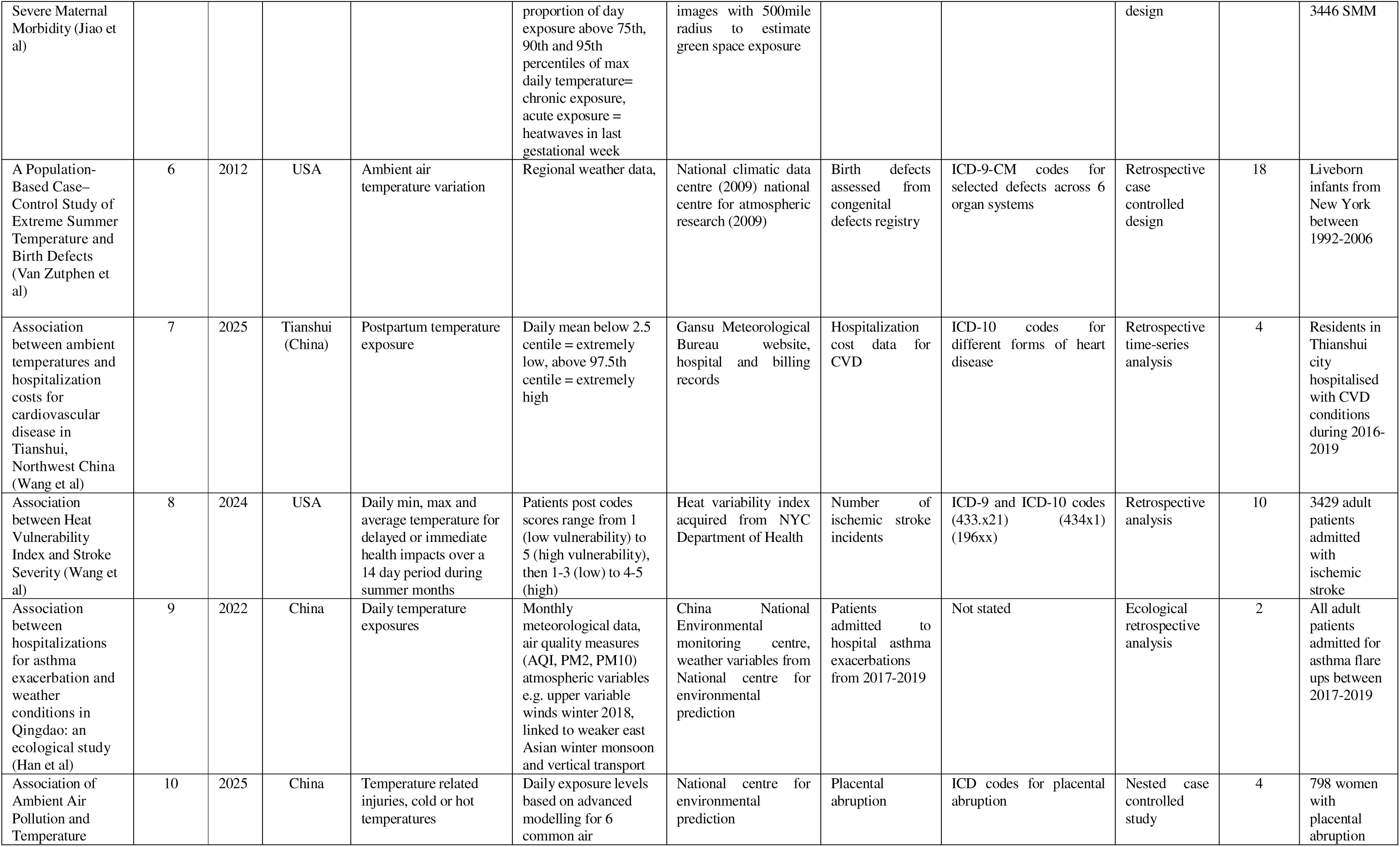
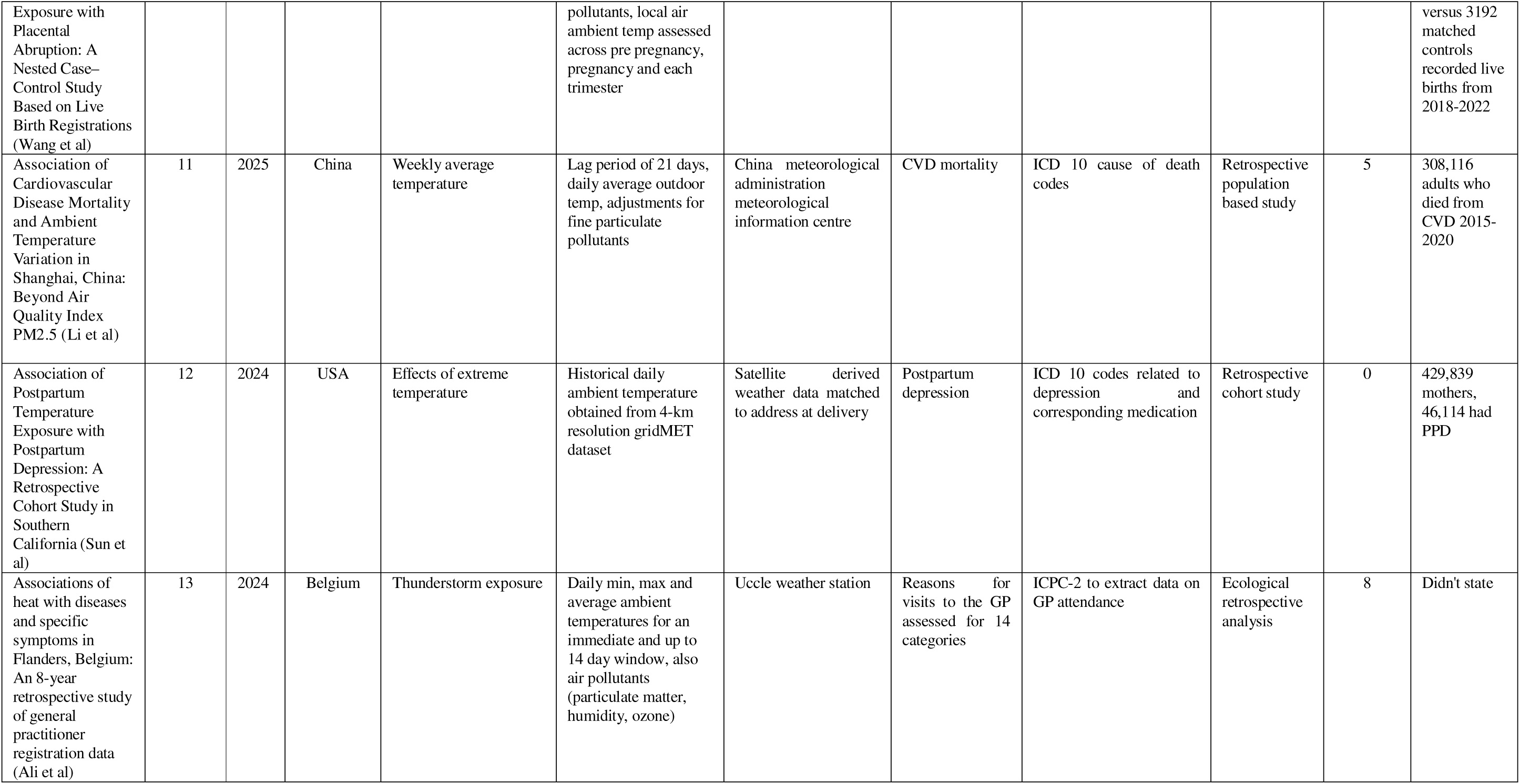
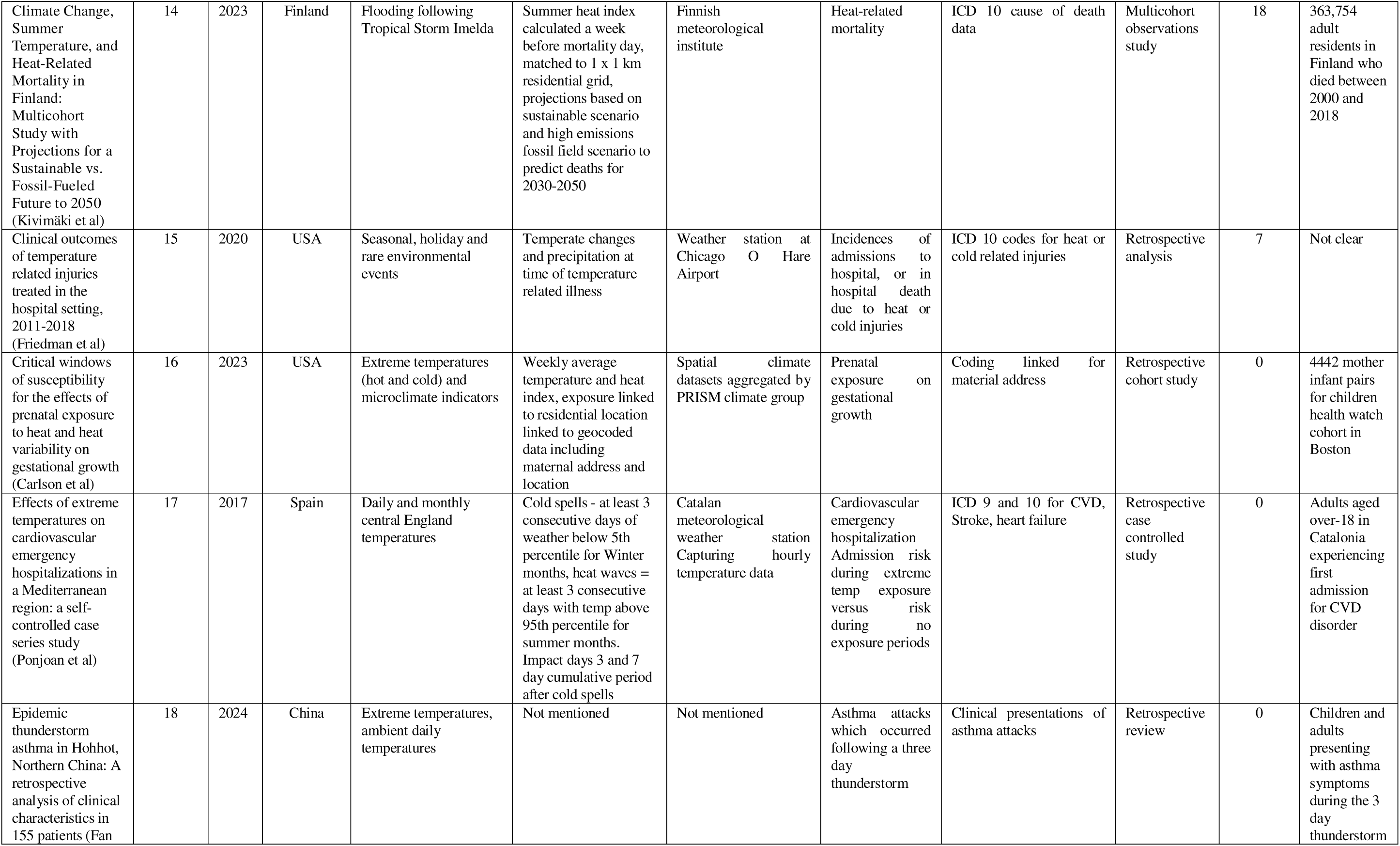
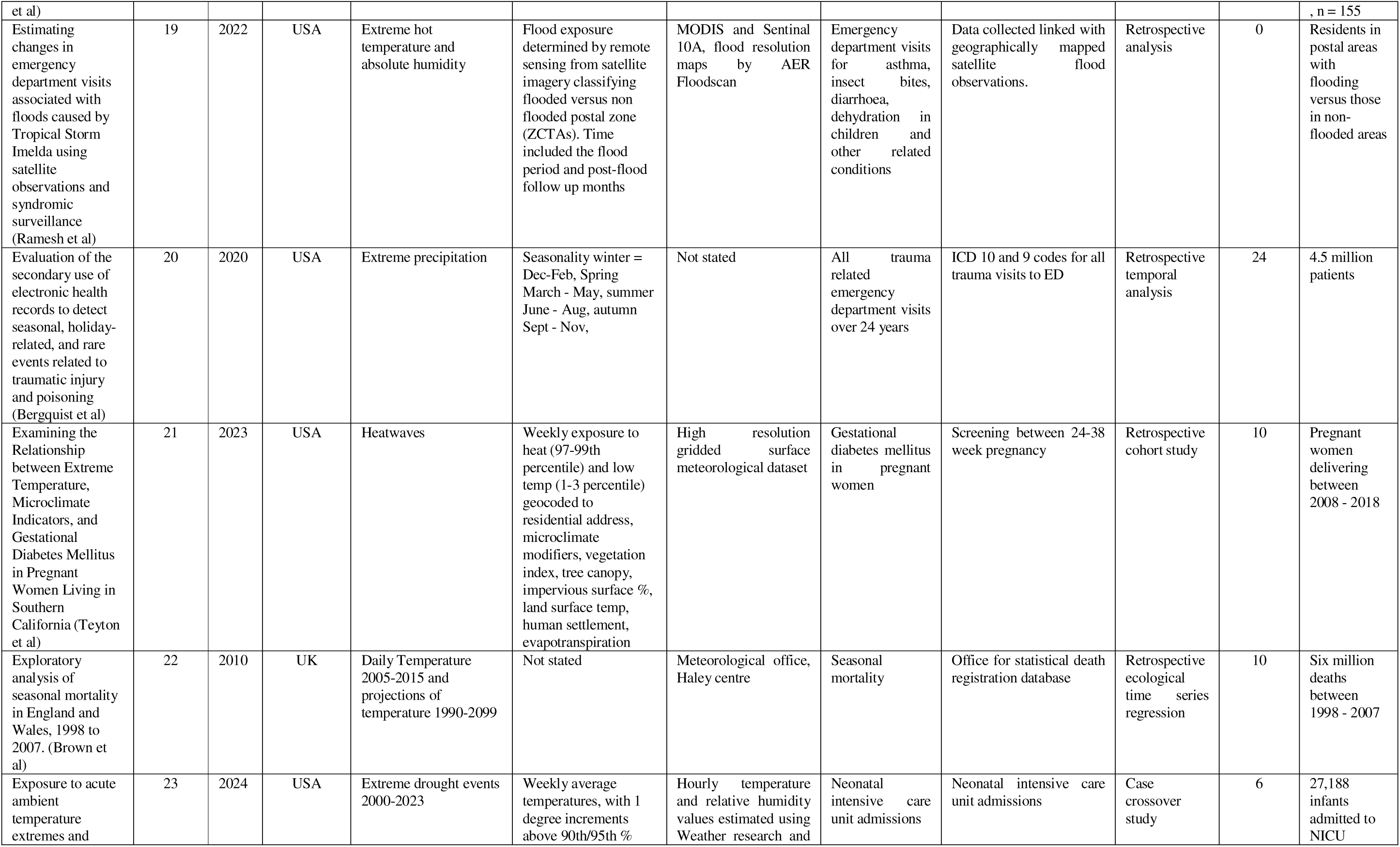
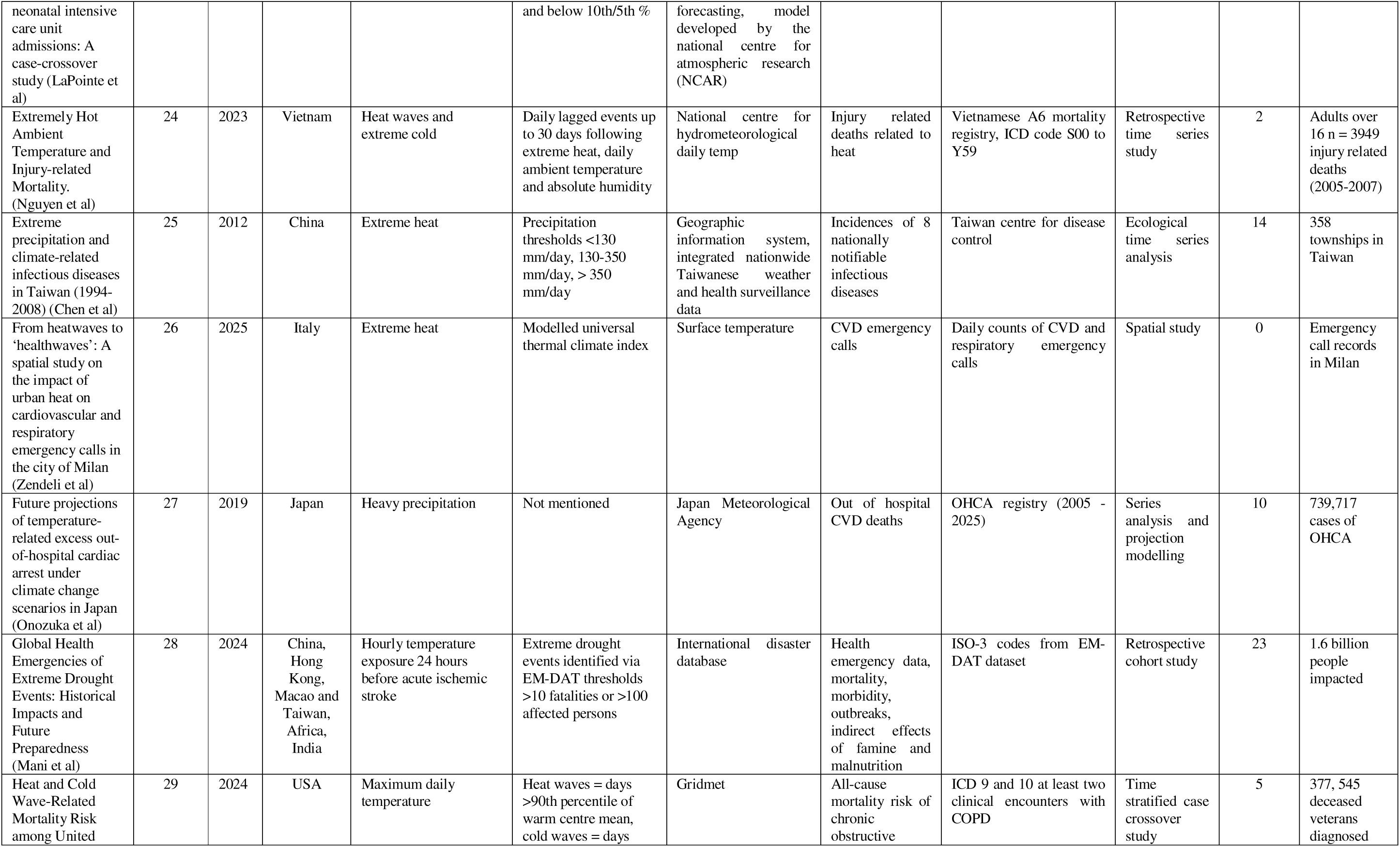
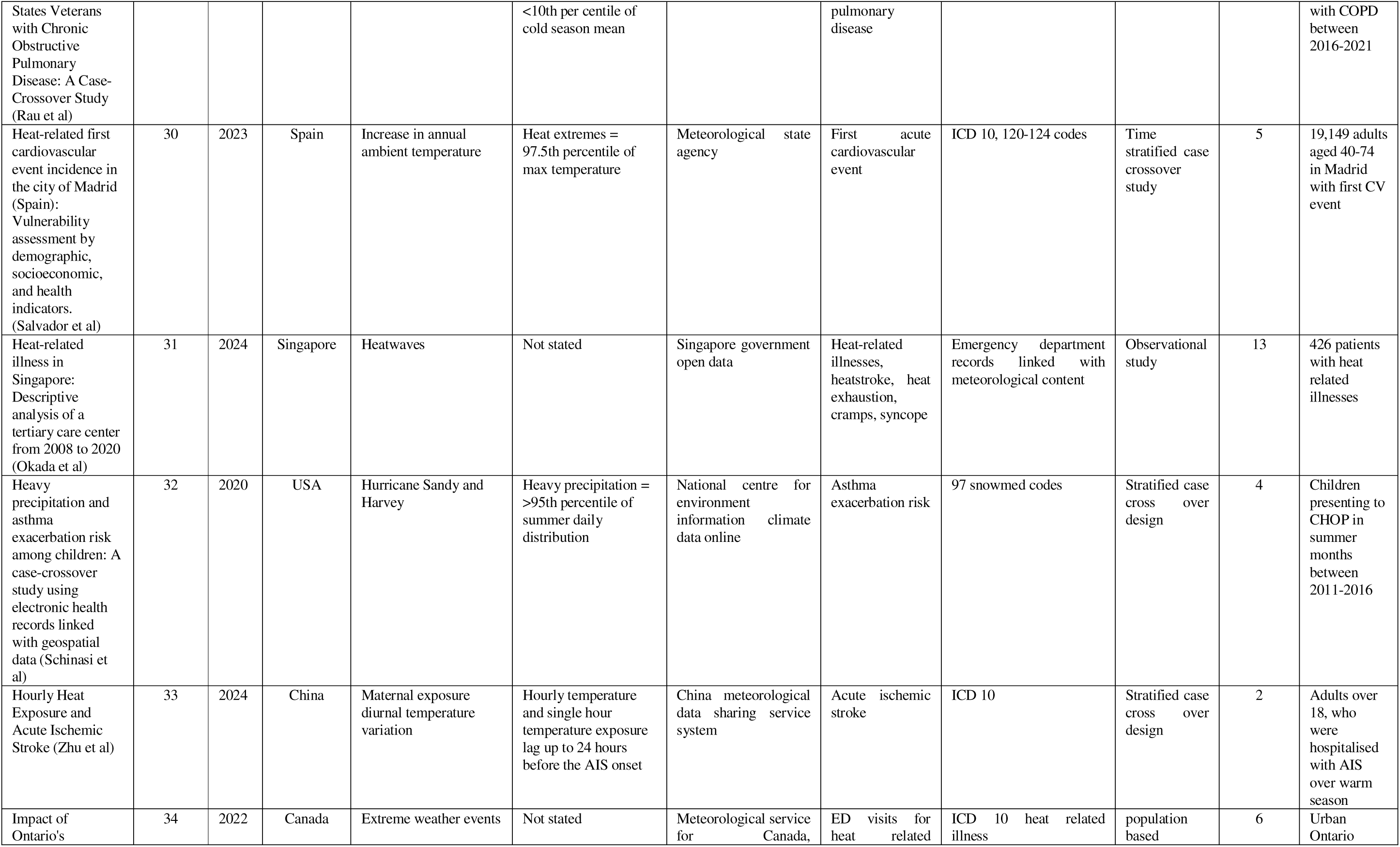
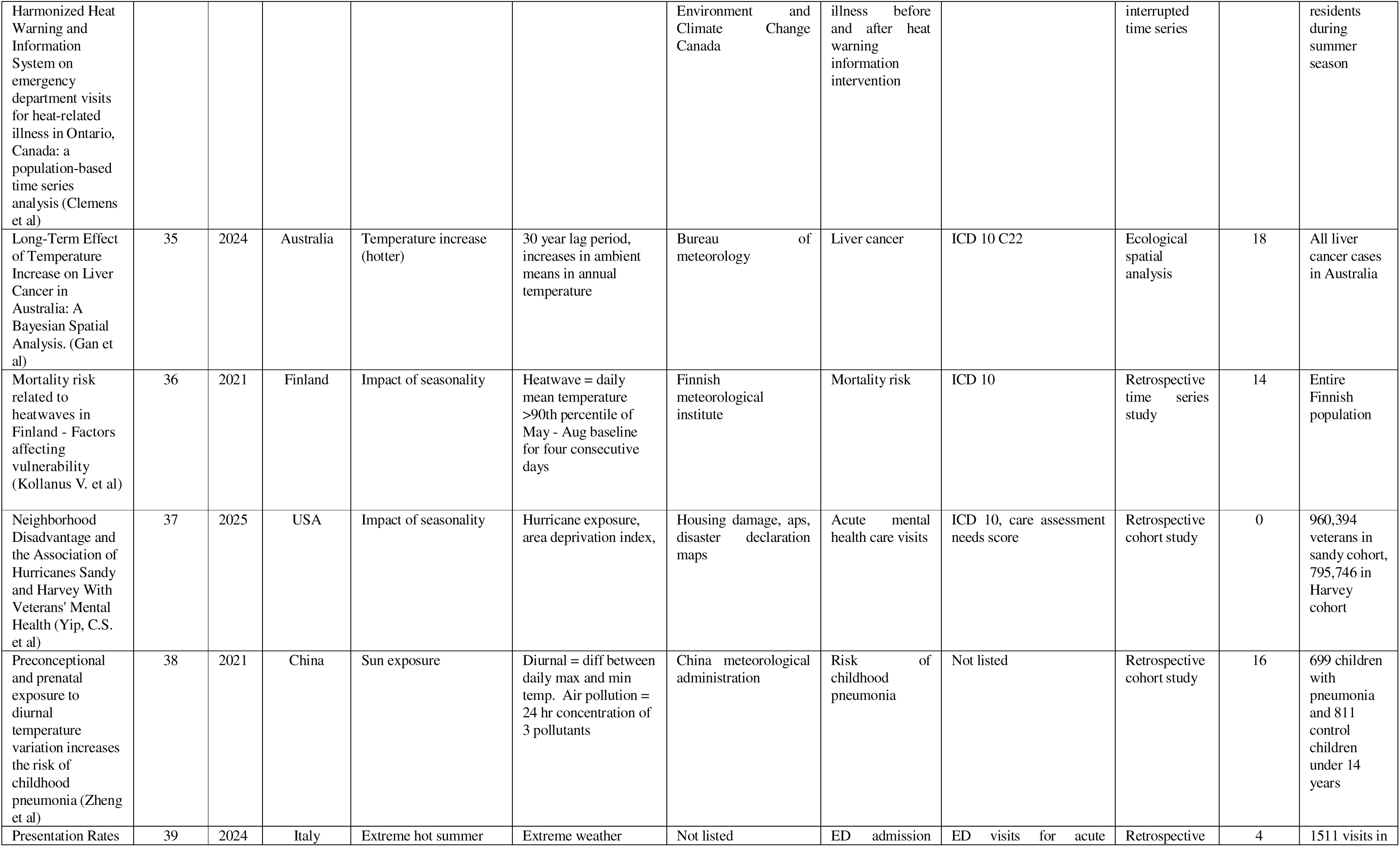
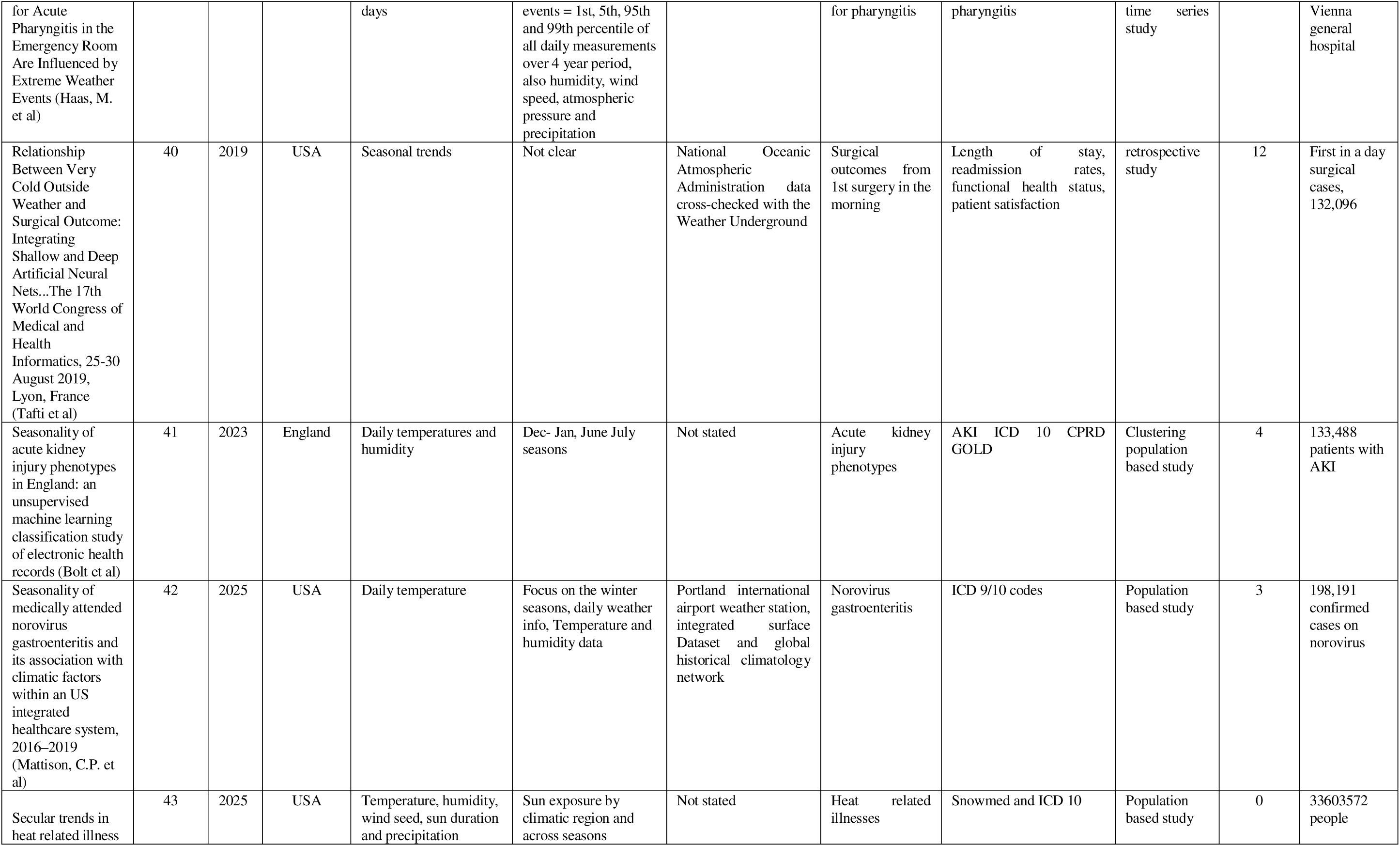
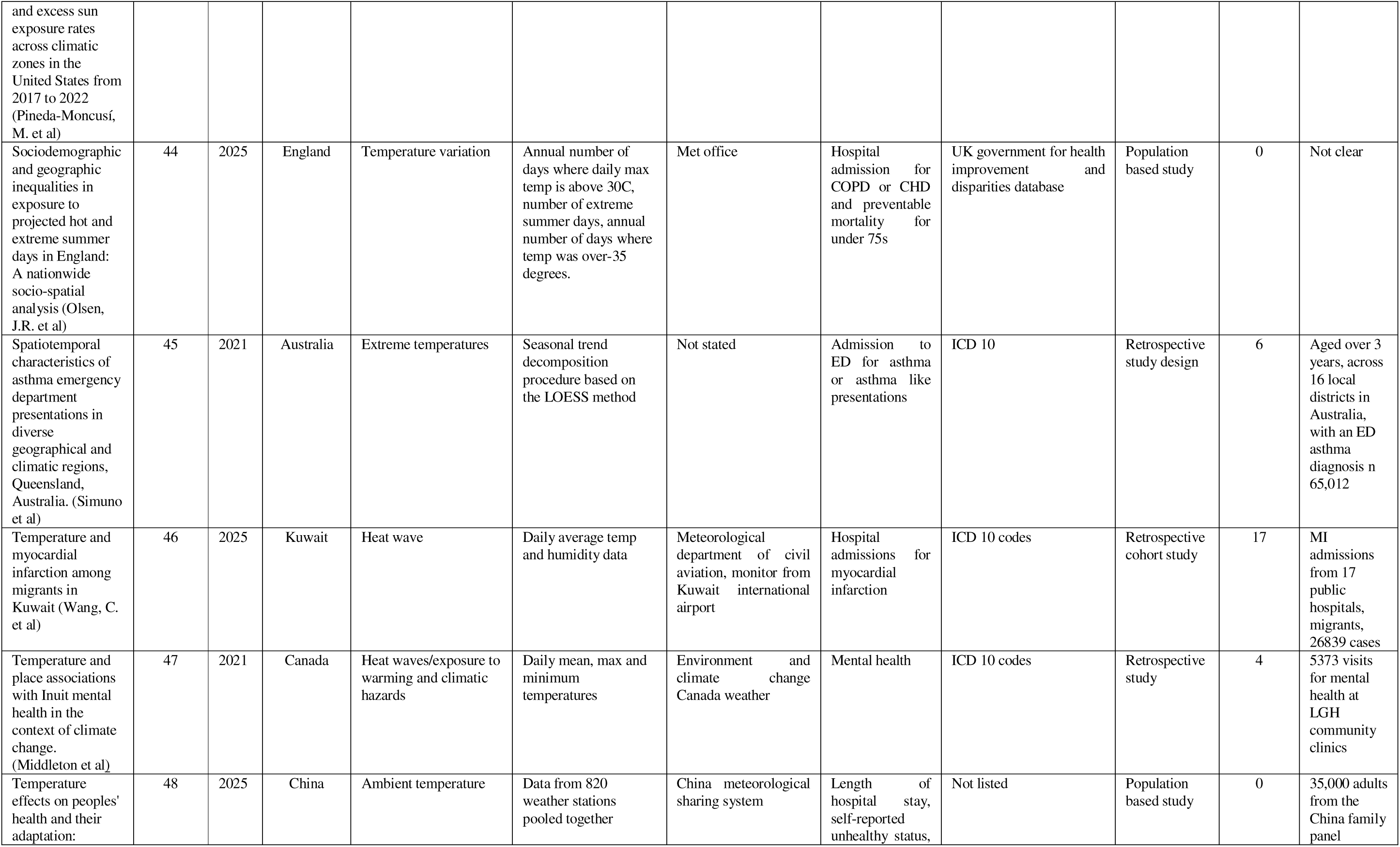
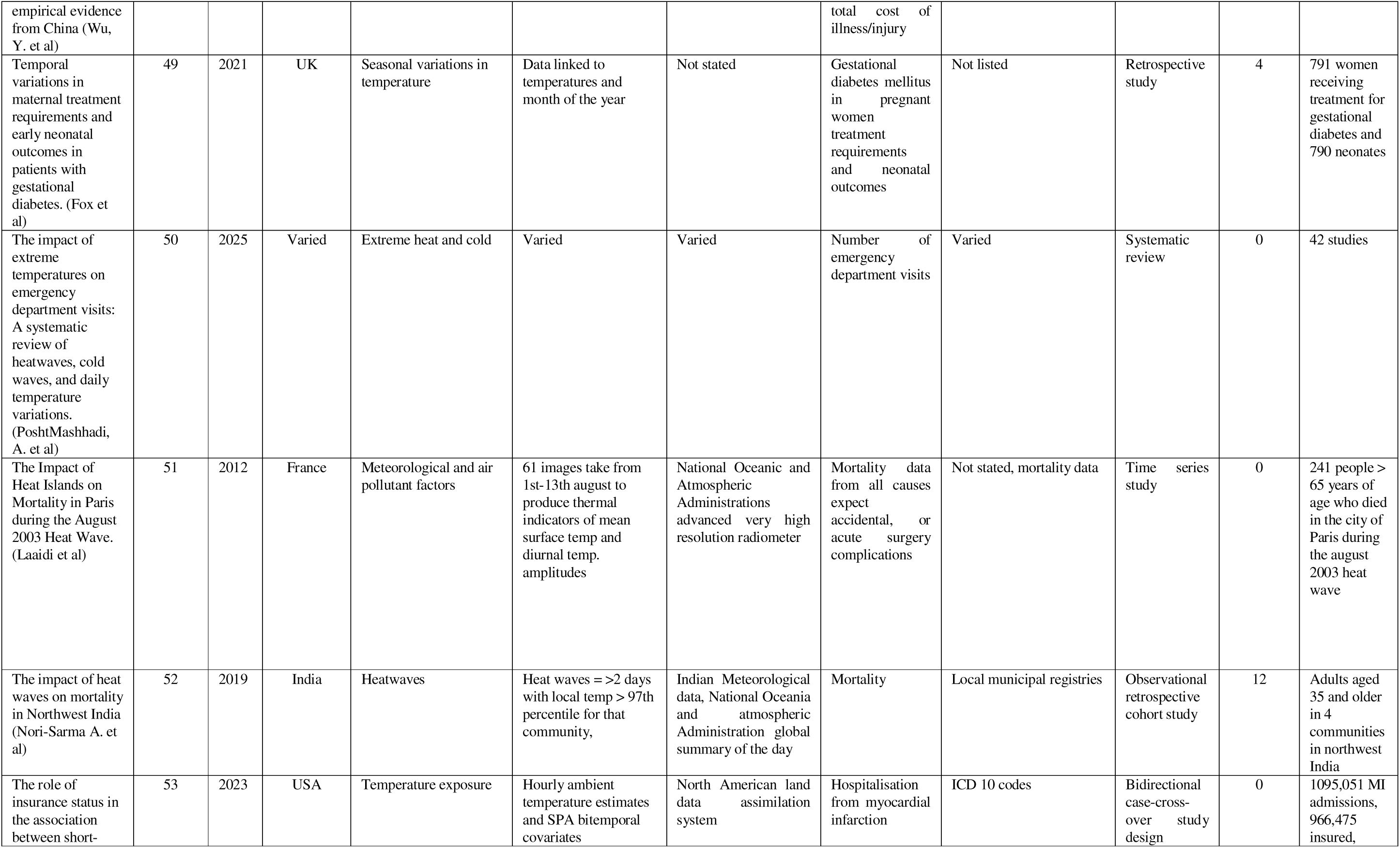
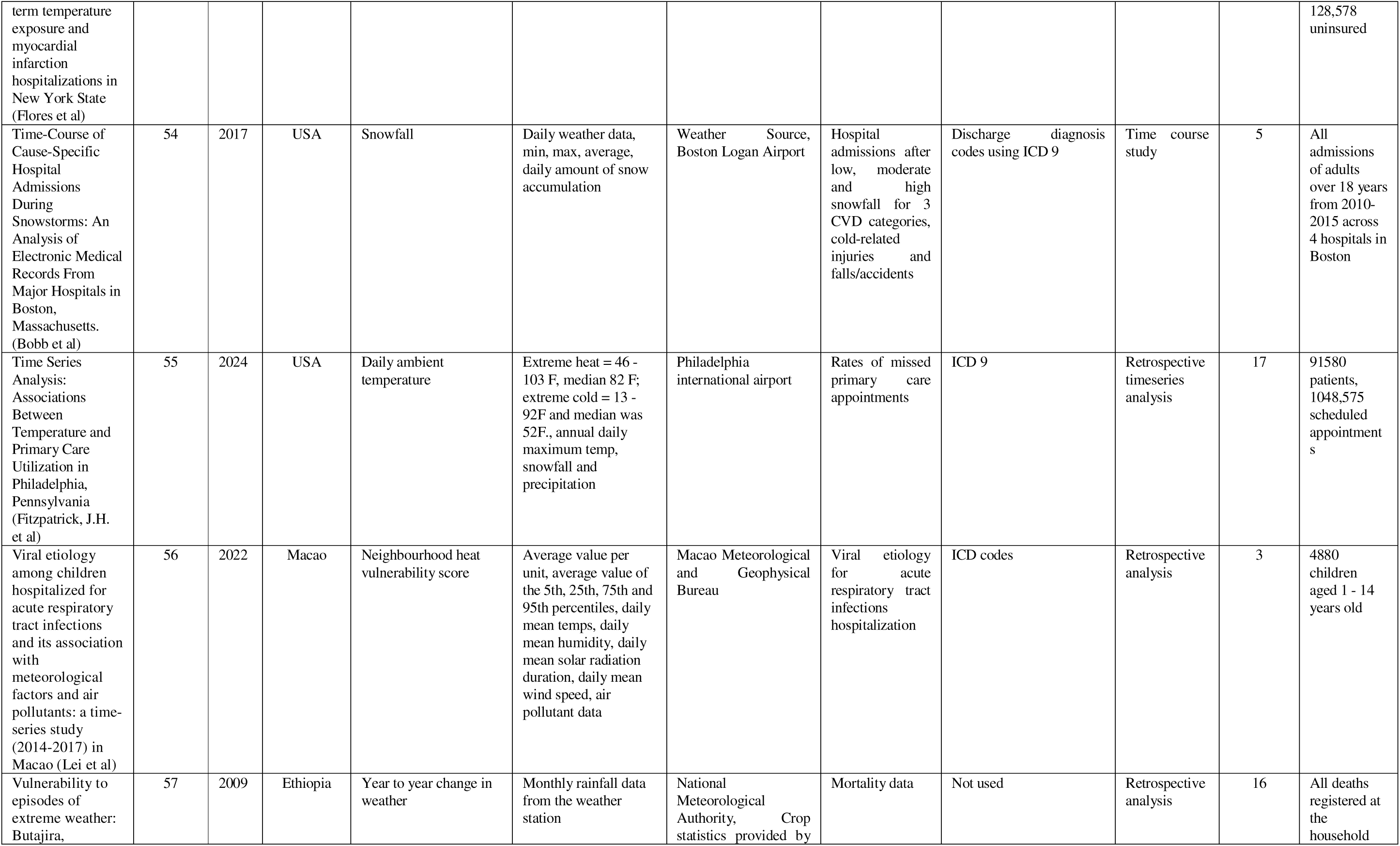
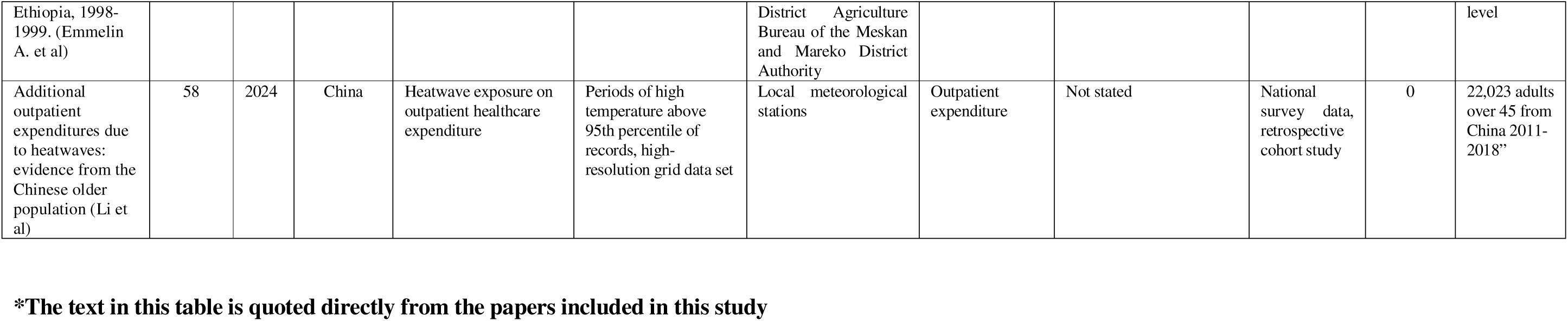
Study characteristics*.

