## Supplementary material for "Examining the Health Impacts of Climate Change Through Electronic Health Records: A Rapid Review"

**Appendix: Supplementary material**

Supplementary Table 1: Search terms and number of papers extracted by database

| **Database** | **Terms Used** | **Results (English)** |
| --- | --- | --- |
| EMBASE | (climate change/ OR climate/)  OR (temperature extreme*.ti,ab. OR heatwave*.ti,ab. OR hot weather.ti,ab. OR cold weather.ti,ab. OR freeze*.ti,ab. OR cold spell*.ti,ab. OR extreme humidity.ti,ab. OR climate.ti,ab. OR extreme weather.ti,ab.)  AND  (electronic health record/  OR electronic health record*.ti,ab.  OR electronic medical record*.ti,ab.  OR routin* collected data.ti,ab.  OR routin* collected health* data.ti,ab.  OR routin*-collected data.ti,ab.  OR health adj2 admin* data.ti,ab.  OR registr* data.ti,ab.) | 55 |
| GREEN FILE | (climate change OR climate OR temperature extreme* OR heatwave* OR hot weather OR cold weather OR freeze* OR cold spell* OR extreme humidity OR extreme weather)  AND  (electronic health record* OR electronic medical record* OR routin* collected data OR routin* collected health* data OR routin*-collected data OR health N2 admin* data OR registr* data) | 26 |
| Web of Science | TS=("climate change" OR climate OR "temperature extreme*" OR heatwave* OR "hot weather" OR "cold weather" OR freeze* OR "cold spell*" OR "extreme humidity" OR “extreme weather”)  AND  TS=("electronic health record*" OR "electronic medical record*" OR "routin* collected data" OR "routin* collected health* data" OR "routin*-collected data" OR "health admin* data" OR "registr* data") | 478 |
| Scopus | TITLE-ABS-KEY(("climate change" OR climate OR "temperature extreme*" OR heatwave* OR "hot weather" OR "cold weather" OR freeze* OR "cold spell*" OR "extreme humidity" OR "extreme weather") AND ("electronic health record*" OR "electronic medical record*" OR "routin* collected data" OR "routin* collected health* data" OR "routin*-collected data" OR "health admin* data" OR "registr* data")) | 436 |
| CINAHL (via EBSCOhost) | (climate change OR climate OR temperature extreme* OR heatwave* OR hot weather OR cold weather OR freeze* OR cold spell* OR extreme humidity)  AND  (electronic health record* OR electronic medical record* OR routin* collected data OR routin* collected health* data OR routin*-collected data OR health N2 admin* data OR registr* data) | 576 |
| MEDLINE (Ovid) | (Climate Change/ OR Climate/)  OR (temperature extreme*.ti,ab. OR heatwave*.ti,ab. OR hot weather.ti,ab. OR cold weather.ti,ab. OR freeze*.ti,ab. OR cold spell*.ti,ab. OR extreme humidity.ti,ab. OR climate.ti,ab.OR extreme weather.ti,ab.)  AND  (Electronic Health Records/ OR Medical Records Systems, Computerized/  OR electronic health record*.ti,ab.  OR electronic medical record*.ti,ab.  OR routin* collected data.ti,ab.  OR routin* collected health* data.ti,ab.  OR routin*-collected data.ti,ab.  OR health adj2 admin* data.ti,ab.  OR registr* data.ti,ab.) | 30 |
| Cochrane | "climate change" OR climate OR "temperature extreme*" OR heatwave* OR "hot weather" OR "cold weather" OR freeze* OR "cold spell*" OR "extreme humidity"  AND  "electronic health record*" OR "electronic medical record*" OR "routin* collected data" OR "routin* collected health* data" OR "routin*-collected data" OR "health admin* data" OR "registr* data" | 14 |

**Supplementary Table 2: Study characteristics***

| **Study Title and Author** | **Paper number** | **Year** | **Country of origin** | **Climate extreme exposure** | **Definition and metric of exposure variable** | **Data source** | **EHR health outcomes** | **Coding** | **Study design** | **Look back period** | **Population** |
| --- | --- | --- | --- | --- | --- | --- | --- | --- | --- | --- | --- |
| “A Difference-in-Differences Approach to Assess the Effect of a Heat Action Plan on Heat-Related Mortality, and Differences in Effectiveness According to Sex, Age, and Socioeconomic Status (Montreal, Quebec). (Benmarhnia et al) | 1 | 2016 | Canada | Summer heat exposure during embryo-foetal critical window | Not stated | Environment Canada meteorological observation stations | Frequency of non-accidental deaths related to heat | Codes not stated | Observational quasi design | 7 | Residents of Montreal who died during 2000-2007 |
| Adverse Weather Conditions can have Negative Effects on Birth Outcomes: Evidence from a birth registry cohort in Tanzania (Castro et al) | 2 | 2014 | north Tanzania | Daily ambient temperature | Effective drought index, Multivariate ENSO Index | Weather stations | Neonatal birth weight, Apgar scores, premature deaths, | not stated | Observational registry based cohort study | 11 | 30,068 singleton term pregnancies, lower risk cohorts |
| Ambient temperature and early delivery of Singleton Pregnancies (Ha et al) | 3 | 2017 | USA | Neighbourhood heat Vulnerability score | Hourly temperature obtained from weather research and forecasting model v3.22 | Weather research and forecasting model v3.22 | Likelihood of early delivery | ICD-9 diagnostic codes, delivery and hospital discharge records | Retrospective observational cohort | 6 | 220,000 singleton births across 12 hospitals |
| Ambient temperature and stillbirth: Risks associated with chronic extreme temperature and acute temperature change. (Kanner et al) | 4 | 2020 | USA | Year to year change in weather | Weather research forecasting model wrf v3.2.1 at a 12 x 12km2 resolution, hourly temperature exposure | Weather research and forecasting model v3.22 | Risk of stillbirth | ICD-9 diagnostic codes, delivery and hospital discharge records | Case controlled study | 0 | 498 still births over 20 weeks gestation |
| Analysis of Heat Exposure During Pregnancy and Severe Maternal Morbidity (Jiao et al) | 5 | 2023 | USA | Ambient air pollution and temperature exposure | Grided meteorological data mapped to maternal residence, proportion of day exposure above 75th, 90th and 95th percentiles of max daily temperature = chronic exposure, acute exposure = heatwaves in last gestational week | 20 condition algorithm; gridMET data, street view images with 500mile radius to estimate green space exposure | Severe maternal morbidity | ICD algorithms refined by CDC, used to classify SSM | Retrospective case controlled design | 10 | 403,602 singleton pregnancies, 3446 SMM |
| A Population-Based Case–Control Study of Extreme Summer Temperature and Birth Defects (Van Zutphen et al) | 6 | 2012 | USA | Ambient air temperature variation | Regional weather data, | National climatic data centre (2009) national centre for atmospheric research (2009) | Birth defects assessed from congenital defects registry | ICD-9-CM codes for selected defects across 6 organ systems | Retrospective case controlled design | 18 | Liveborn infants from New York between 1992-2006 |
| Association between ambient temperatures and hospitalization costs for cardiovascular disease in Tianshui, Northwest China  (Wang et al) | 7 | 2025 | Tianshui (China) | Postpartum temperature exposure | Daily mean below 2.5 centile = extremely low, above 97.5th centile = extremely high | Gansu Meteorological Bureau website, hospital and billing records | Hospitalisation cost data for CVD | ICD-10 codes for different forms of heart disease | Retrospective time-series analysis | 4 | Residents in Thianshui city hospitalised with CVD conditions during 2016-2019 |
| Association between Heat Vulnerability Index and Stroke Severity (Wang et al) | 8 | 2024 | USA | Daily min, max and average temperature for delayed or immediate health impacts over a 14 day period during summer months | Patients post codes scores range from 1 (low vulnerability) to 5 (high vulnerability), then 1-3 (low) to 4-5 (high) | Heat variability index acquired from NYC Department of Health | Number of ischemic stroke incidents | ICD-9 and ICD-10 codes (433.x21) (434x1) (196xx) | Retrospective analysis | 10 | 3429 adult patients admitted with ischemic stroke |
| Association between hospitalizations for asthma exacerbation and weather conditions in Qingdao: an ecological study  (Han et al) | 9 | 2022 | China | Daily temperature exposures | Monthly meteorological data, air quality measures (AQI, PM2, PM10) atmospheric variables e.g. upper variable winds winter 2018, linked to weaker east Asian winter monsoon and vertical transport | China National Environmental monitoring centre, weather variables from National centre for environmental prediction | Patients admitted to hospital asthma exacerbations from 2017-2019 | Not stated | Ecological retrospective analysis | 2 | All adult patients admitted for asthma flare ups between 2017-2019 |
| Association of Ambient Air Pollution and Temperature Exposure with Placental Abruption: A Nested Case–Control Study Based on Live Birth Registrations (Wang et al) | 10 | 2025 | China | Temperature related injuries, cold or hot temperatures | Daily exposure levels based on advanced modelling for 6 common air pollutants, local air ambient temp assessed across pre pregnancy, pregnancy and each trimester | National centre for environmental prediction | Placental abruption | ICD codes for placental abruption | Nested case controlled study | 4 | 798 women with placental abruption versus 3192 matched controls recorded live births from 2018-2022 |
| Association of Cardiovascular Disease Mortality and Ambient Temperature Variation in Shanghai, China: Beyond Air Quality Index PM2.5 (Li et al) | 11 | 2025 | China | Weekly average temperature | Lag period of 21 days, daily average outdoor temp, adjustments for fine particulate pollutants | China meteorological administration meteorological information centre | CVD mortality | ICD 10 cause of death codes | Retrospective population based study | 5 | 308,116 adults who died from CVD 2015-2020 |
| Association of Postpartum Temperature Exposure with Postpartum Depression: A Retrospective Cohort Study in Southern California (Sun et al) | 12 | 2024 | USA | Effects of extreme temperature | Historical daily ambient temperature obtained from 4-km resolution gridMET dataset | Satellite derived weather data matched to address at delivery | Postpartum depression | ICD 10 codes related to depression and corresponding medication | Retrospective cohort study | 0 | 429,839 mothers, 46,114 had PPD |
| Associations of heat with diseases and specific symptoms in Flanders, Belgium: An 8-year retrospective study of general practitioner registration data (Ali et al) | 13 | 2024 | Belgium | Thunderstorm exposure | Daily min, max and average ambient temperatures for an immediate and up to 14 day window, also air pollutants (particulate matter, humidity, ozone) | Uccle weather station | Reasons for visits to the GP assessed for 14 categories | ICPC-2 to extract data on GP attendance | Ecological retrospective analysis | 8 | Didn't state |
| Climate Change, Summer Temperature, and Heat-Related Mortality in Finland: Multicohort Study with Projections for a Sustainable vs. Fossil-Fueled Future to 2050  (Kivimäki et al) | 14 | 2023 | Finland | Flooding following Tropical Storm Imelda | Summer heat index calculated a week before mortality day, matched to 1 x 1 km residential grid, projections based on sustainable scenario and high emissions fossil field scenario to predict deaths for 2030-2050 | Finnish meteorological institute | Heat-related mortality | ICD 10 cause of death data | Multicohort observations study | 18 | 363,754 adult residents in Finland who died between 2000 and 2018 |
| Clinical outcomes of temperature related injuries treated in the hospital setting, 2011-2018 (Friedman et al) | 15 | 2020 | USA | Seasonal, holiday and rare environmental events | Temperate changes and precipitation at time of temperature related illness | Weather station at Chicago O Hare Airport | Incidences of admissions to hospital, or in hospital death due to heat or cold injuries | ICD 10 codes for heat or cold related injuries | Retrospective analysis | 7 | Not clear |
| Critical windows of susceptibility for the effects of prenatal exposure to heat and heat variability on gestational growth (Carlson et al) | 16 | 2023 | USA | Extreme temperatures (hot and cold) and microclimate indicators | Weekly average temperature and heat index, exposure linked to residential location linked to geocoded data including maternal address and location | Spatial climate datasets aggregated by PRISM climate group | Prenatal exposure on gestational growth | Coding linked for material address | Retrospective cohort study | 0 | 4442 mother infant pairs for children health watch cohort in Boston |
| Effects of extreme temperatures on cardiovascular emergency hospitalizations in a Mediterranean region: a self-controlled case series study (Ponjoan et al) | 17 | 2017 | Spain | Daily and monthly central England temperatures | Cold spells - at least 3 consecutive days of weather below 5th percentile for Winter months, heat waves = at least 3 consecutive days with temp above 95th percentile for summer months. Impact days 3 and 7 day cumulative period after cold spells | Catalan meteorological weather station Capturing hourly temperature data | Cardiovascular emergency hospitalisation Admission risk during extreme temp exposure versus risk during no exposure periods | ICD 9 and 10 for CVD, Stroke, heart failure | Retrospective case controlled study | 0 | Adults aged over-18 in Catalonia experiencing first admission for CVD disorder |
| Epidemic thunderstorm asthma in Hohhot, Northern China: A retrospective analysis of clinical characteristics in 155 patients (Fan et al) | 18 | 2024 | China | Extreme temperatures, ambient daily temperatures | Not mentioned | Not mentioned | Asthma attacks which occurred following a three day thunderstorm | Clinical presentations of asthma attacks | Retrospective review | 0 | Children and adults presenting with asthma symptoms during the 3 day thunderstorm, n = 155 |
| Estimating changes in emergency department visits associated with floods caused by Tropical Storm Imelda using satellite observations and syndromic surveillance (Ramesh et al) | 19 | 2022 | USA | Extreme hot temperature and absolute humidity | Flood exposure determined by remote sensing from satellite imagery classifying flooded versus non flooded postal zone (ZCTAs). Time included the flood period and post-flood follow up months | MODIS and Sentinal 10A, flood resolution maps by AER Floodscan | Emergency department visits for asthma, insect bites, diarrhoea, dehydration in children and other related conditions | Data collected linked with geographically mapped satellite flood observations. | Retrospective analysis | 0 | Residents in postal areas with flooding versus those in non-flooded areas |
| Evaluation of the secondary use of electronic health records to detect seasonal, holiday-related, and rare events related to traumatic injury and poisoning (Bergquist et al) | 20 | 2020 | USA | Extreme precipitation | Seasonality winter = dec-feb, Spring March - May, summer June - Aug, autumn Sept - Nov, | Not stated | All trauma related emergency department visits over 24 years | ICD 10 and 9 codes for all trauma visits to ED | Retrospective temporal analysis | 24 | 4.5 million patients |
| Examining the Relationship between Extreme Temperature, Microclimate Indicators, and Gestational Diabetes Mellitus in Pregnant Women Living in Southern California (Teyton et al) | 21 | 2023 | USA | Heatwaves | Weekly exposure to heat (97-99th percentile) and low temp (1-3 percentile) geocoded to residential address, microclimate modifiers, vegetation index, tree canopy, impervious surface %, land surface temp, human settlement, evapotranspiration | High resolution gridded surface meteorological dataset | Gestational diabetes mellitus in pregnant women | Screening between 24-38 week pregnancy | Retrospective cohort study | 10 | Pregnant women delivering between 2008 - 2018 |
| Exploratory analysis of seasonal mortality in England and Wales, 1998 to 2007. (Brown et al) | 22 | 2010 | UK | Daily Temperature 2005-2015 and projections of temperature 1990-2099 | Not stated | Meteorological office, Haley centre | Seasonal mortality | Office for statistical death registration database | Retrospective ecological time series regression | 10 | Six million deaths between 1998 - 2007 |
| Exposure to acute ambient temperature extremes and neonatal intensive care unit admissions: A case-crossover study (LaPointe et al) | 23 | 2024 | USA | Extreme drought events 2000-2023 | Weekly average temperatures, with 1 degree increments above 90th/95th % and below 10th/5th % | Hourly temperature and relative humidity values estimated using Weather research and forecasting, model developed by the national centre for atmospheric research (NCAR) | Neonatal intensive care unit admissions | Neonatal intensive care unit admissions | Case crossover study | 6 | 27,188 infants admitted to NICU |
| Extremely Hot Ambient Temperature and Injury-related Mortality. (Nguyen et al) | 24 | 2023 | Vietnam | Heat waves and extreme cold | Daily lagged events up to 30 days following extreme heat, daily ambient temperature and absolute humidity | National centre for hydrometeorological daily temp | Injury related deaths related to heat | Vietnamese A6 mortality registry, ICD code S00 to Y59 | Retrospective time series study | 2 | Adults over 16 n = 3949 injury related deaths (2005-2007) |
| Extreme precipitation and climate-related infectious diseases in Taiwan (1994-2008) (Chen et al) | 25 | 2012 | China | Extreme heat | Precipitation thresholds <130 mm/day, 130-350 mm/day, > 350 mm/day | Geographic information system, integrated nationwide Taiwanese weather and health surveillance data | Incidences of 8 nationally notifiable infectious diseases | Taiwan centre for disease control | Ecological time series analysis | 14 | 358 townships in Taiwan |
| From heatwaves to ‘healthwaves’: A spatial study on the impact of urban heat on cardiovascular and respiratory emergency calls in the city of Milan (Zendeli et al) | 26 | 2025 | Italy | Extreme heat | Modelled universal thermal climate index | Surface temperature | CVD emergency calls | Daily counts of CVD and respiratory emergency calls | Spatial study | 0 | Emergency call records in Milan |
| Future projections of temperature-related excess out-of-hospital cardiac arrest under climate change scenarios in Japan (Onozuka et al) | 27 | 2019 | Japan | Heavy precipitation | Not mentioned | Japan Meteorological Agency | Out of hospital CVD deaths | OHCA registry (2005 - 2025) | Series analysis and projection modelling | 10 | 739,717 cases of OHCA |
| Global Health Emergencies of Extreme Drought Events: Historical Impacts and Future Preparedness (Mani et al) | 28 | 2024 | China, Hong Kong, Macao and Taiwan, Africa, India | Hourly temperature exposure 24 hours before acute ischemic stroke | Extreme drought events identified via EM-DAT thresholds >10 fatalities or >100 affected persons | International disaster database | Health emergency data, mortality, morbidity, outbreaks, indirect effects of famine and malnutrition | ISO-3 codes from EM-DAT dataset | Retrospective cohort study | 23 | 1.6 billion people impacted |
| Heat and Cold Wave-Related Mortality Risk among United States Veterans with Chronic Obstructive Pulmonary Disease: A Case-Crossover Study (Rau et al) | 29 | 2024 | USA | Maximum daily temperature | Heat waves = days >90th percentile of warm centre mean, cold waves = days <10th per centile of cold season mean | Gridmet | All-cause mortality risk of chronic obstructive pulmonary disease | ICD 9 and 10 at least two clinical encounters with COPD | Time stratified case crossover study | 5 | 377, 545 deceased veterans diagnosed with COPD between 2016-2021 |
| Heat-related first cardiovascular event incidence in the city of Madrid (Spain): Vulnerability assessment by demographic, socioeconomic, and health indicators. (Salvador et al) | 30 | 2023 | Spain | Increase in annual ambient temperature | Heat extremes = 97.5th percentile of max temperature | Meteorological state agency | First acute cardiovascular event | ICD 10, 120-124 codes | Time stratified case crossover study | 5 | 19,149 adults aged 40-74 in Madrid with first CV event |
| Heat-related illness in Singapore: Descriptive analysis of a tertiary care center from 2008 to 2020 (Okada et al) | 31 | 2024 | Singapore | Heatwaves | Not stated | Singapore government open data | Heat-related illnesses, heatstroke, heat exhaustion, cramps, syncope | Emergency department records linked with meteorological content | Observational study | 13 | 426 patients with heat related illnesses |
| Heavy precipitation and asthma exacerbation risk among children: A case-crossover study using electronic health records linked with geospatial data (Schinasi et al) | 32 | 2020 | USA | Hurricane Sandy and Harvey | Heavy precipitation = >95th percentile of summer daily distribution | National centre for environment information climate data online | Asthma exacerbation risk | 97 snowmed codes | Stratified case cross over design | 4 | Children presenting to CHOP in summer months between 2011-2016 |
| Hourly Heat Exposure and Acute Ischemic Stroke (Zhu et al) | 33 | 2024 | China | Maternal exposure diurnal temperature variation | Hourly temperature and single hour temperature exposure lag up to 24 hours before the AIS onset | China meteorological data sharing service system | Acute ischemic stroke | ICD 10 | Stratified case cross over design | 2 | Adults over 18, who were hospitalised with AIS over warm season |
| Impact of Ontario's Harmonized Heat Warning and Information System on emergency department visits for heat-related illness in Ontario, Canada: a population-based time series analysis (Clemens et al) | 34 | 2022 | Canada | Extreme weather events | Not stated | Meteorological service for Canada, Environment and Climate Change Canada | ED visits for heat related illness before and after heat warning information intervention | ICD 10 heat related illness | population based interrupted time series | 6 | Urban Ontario residents during summer season |
| Long-Term Effect of Temperature Increase on Liver Cancer in Australia: A Bayesian Spatial Analysis. (Gan et al) | 35 | 2024 | Australia | Temperature increase (hotter) | 30 year lag period, increases in ambient means in annual temperature | Bureau of meteorology | Liver cancer | ICD 10 C22 | Ecological spatial analysis | 18 | All liver cancer cases in Australia |
| Mortality risk related to heatwaves in Finland - Factors affecting vulnerability (Kollanus V. et al) | 36 | 2021 | Finland | Impact of seasonality | Heatwave = daily mean temperature >90th percentile of May - Aug baseline for four consecutive days | Finnish meteorological institute | Mortality risk | ICD 10 | Retrospective time series study | 14 | Entire Finnish population |
| Neighborhood Disadvantage and the Association of Hurricanes Sandy and Harvey With Veterans' Mental Health (Yip, C.S. et al) | 37 | 2025 | USA | Impact of seasonality | Hurricane exposure, area deprivation index, | Housing damage, aps, disaster declaration maps | Acute mental health care visits | ICD 10, care assessment needs score | Retrospective cohort study | 0 | 960,394 veterans in sandy cohort, 795,746 in Harvey cohort |
| Preconceptional and prenatal exposure to diurnal temperature variation increases the risk of childhood pneumonia (Zheng et al) | 38 | 2021 | China | Sun exposure | Diurnal = diff between daily max and min temp. Air pollution = 24 hr concentration of 3 pollutants | China meteorological administration | Risk of childhood pneumonia | Not listed | Retrospective cohort study | 16 | 699 children with pneumonia and 811 control children under 14 years |
| Presentation Rates for Acute Pharyngitis in the Emergency Room Are Influenced by Extreme Weather Events (Haas, M. et al) | 39 | 2024 | Italy | Extreme hot summer days | Extreme weather events = 1st, 5th, 95th and 99th percentile of all daily measurements over 4 year period, also humidity, wind speed, atmospheric pressure and precipitation | Not listed | ED admission for pharyngitis | ED visits for acute pharyngitis | Retrospective time series study | 4 | 1511 visits in Vienna general hospital |
| Relationship Between Very Cold Outside Weather and Surgical Outcome: Integrating Shallow and Deep Artificial Neural Nets...The 17th World Congress of Medical and Health Informatics, 25-30 August 2019, Lyon, France (Tafti et al) | 40 | 2019 | USA | Seasonal trends | Not clear | National Oceanic Atmospheric Administration data cross-checked with the Weather Underground | Surgical outcomes from 1st surgery in the morning | Length of stay, readmission rates, functional health status, patient satisfaction | retrospective study | 12 | First in a day surgical cases, 132,096 |
| Seasonality of acute kidney injury phenotypes in England: an unsupervised machine learning classification study of electronic health records (Bolt et al) | 41 | 2023 | England | Daily temperatures and humidity | Dec- Jan, June July seasons | Not stated | Acute kidney injury phenotypes | AKI ICD 10 CPRD GOLD | Clustering population based study | 4 | 133,488 patients with AKI |
| Seasonality of medically attended norovirus gastroenteritis and its association with climatic factors within an US integrated healthcare system, 2016–2019 (Mattison, C.P. et al) | 42 | 2025 | USA | Daily temperature | Focus on the winter seasons, daily weather info, Temperature and humidity data | Portland international airport weather station, integrated surface Dataset and global historical climatology network | Norovirus gastroenteritis | ICD 9/10 codes | Population based study | 3 | 198,191 confirmed cases on norovirus |
| Secular trends in heat related illness and excess sun exposure rates across climatic zones in the United States from 2017 to 2022 (Pineda-Moncusí, M. et al) | 43 | 2025 | USA | Temperature, humidity, wind seed, sun duration and precipitation | Sun exposure by climatic region and across seasons | Not stated | Heat related illnesses | Snowmed and ICD 10 | Population based study | 0 | 33603572 people |
| Sociodemographic and geographic inequalities in exposure to projected hot and extreme summer days in England: A nationwide socio-spatial analysis (Olsen, J.R. et al) | 44 | 2025 | England | Temperature variation | Annual number of days where daily max temp is above 30C, number of extreme summer days, annual number of days where temp was over-35 degrees. | Met office | Hospital admission for COPD or CHD and preventable mortality for under 75s | UK government for health improvement and disparities database | Population based study | 0 | Not clear |
| Spatiotemporal characteristics of asthma emergency department presentations in diverse geographical and climatic regions, Queensland, Australia. (Simuno et al) | 45 | 2021 | Australia | Extreme temperatures | Seasonal trend decomposition procedure based on the LOESS method | Not stated | Admission to ED for asthma or asthma like presentations | ICD 10 | Retrospective study design | 6 | Aged over 3 years, across 16 local districts in Australia, with an ED asthma diagnosis n 65,012 |
| Temperature and myocardial infarction among migrants in Kuwait (Wang, C. et al) | 46 | 2025 | Kuwait | Heat wave | Daily average temp and humidity data | Meteorological department of civil aviation, monitor from Kuwait international airport | Hospital admissions for myocardial infarction | ICD 10 codes | Retrospective cohort study | 17 | MI admissions from 17 public hospitals, migrants, 26839 cases |
| Temperature and place associations with Inuit mental health in the context of climate change. (Middleton et al) | 47 | 2021 | Canada | Heat waves/exposure to warming and climatic hazards | Daily mean, max and minimum temperatures | Environment and climate change Canada weather | Mental health | ICD 10 codes | Retrospective study | 4 | 5373 visits for mental health at LGH community clinics |
| Temperature effects on peoples' health and their adaptation: empirical evidence from China (Wu, Y. et al) | 48 | 2025 | China | Ambient temperature | Data from 820 weather stations pooled together | China meteorological sharing system | Length of hospital stay, self-reported unhealthy status, total cost of illness/injury | Not listed | Population based study | 0 | 35,000 adults from the China family panel |
| Temporal variations in maternal treatment requirements and early neonatal outcomes in patients with gestational diabetes. (Fox et al) | 49 | 2021 | UK | Seasonal variations in temperature | Data linked to temperatures and month of the year | Not stated | Gestational diabetes mellitus in pregnant women treatment requirements and neonatal outcomes | Not listed | Retrospective study | 4 | 791 women receiving treatment for gestational diabetes and 790 neonates |
| The impact of extreme temperatures on emergency department visits: A systematic review of heatwaves, cold waves, and daily temperature variations. (PoshtMashhadi, A. et al) | 50 | 2025 | Varied | Extreme heat and cold | Varied | Varied | Number of emergency department visits | Varied | Systematic review | 0 | 42 studies |
| The Impact of Heat Islands on Mortality in Paris during the August 2003 Heat Wave. (Laaidi et al) | 51 | 2012 | France | Meteorological and air pollutant factors | 61 images take from 1st-13th august to produce thermal indicators of mean surface temp and diurnal temp. amplitudes | National Oceanic and Atmospheric Administrations advanced very high resolution radiometer | Mortality data from all causes expect accidental, or acute surgery complications | Not stated, mortality data | Time series study | 0 | 241 people > 65 years of age who died in the city of Paris during the august 2003 heat wave |
| The impact of heat waves on mortality in Northwest India (Nori-Sarma A. et al) | 52 | 2019 | India | Heatwaves | Heat waves = >2 days with local temp > 97th percentile for that community, | Indian Meteorological data, National Oceania and atmospheric Administration global summary of the day | Mortality | Local municipal registries | Observational retrospective cohort study | 12 | Adults aged 35 and older in 4 communities in northwest India |
| The role of insurance status in the association between short-term temperature exposure and myocardial infarction hospitalizations in New York State (Flores et al) | 53 | 2023 | USA | Temperature exposure | Hourly ambient temperature estimates and SPA bitemporal covariates | North American land data assimilation system | Hospitalisation from myocardial infarction | ICD 10 codes | Bidirectional case-cross-over study design | 0 | 1095,051 MI admissions, 966,475 insured, 128,578 uninsured |
| Time-Course of Cause-Specific Hospital Admissions During Snowstorms: An Analysis of Electronic Medical Records From Major Hospitals in Boston, Massachusetts. (Bobb et al) | 54 | 2017 | USA | Snowfall | Daily weather data, min, max, average, daily amount of snow accumulation | Weather Source, Boston Logan Airport | Hospital admissions after low, moderate and high snowfall for 3 CVD categories, cold-related injuries and falls/accidents | Discharge diagnosis codes using ICD 9 | Time course study | 5 | All admissions of adults over 18 years from 2010-2015 across 4 hospitals in Boston |
| Time Series Analysis: Associations Between Temperature and Primary Care Utilization in Philadelphia, Pennsylvania (Fitzpatrick, J.H. et al) | 55 | 2024 | USA | Daily ambient temperature | Extreme heat = 46 - 103 F, median 82 F; extreme cold = 13 - 92F and median was 52F., annual daily maximum temp, snowfall and precipitation | Philadelphia international airport | Rates of missed primary care appointments | ICD 9 | Retrospective timeseries analysis | 17 | 91580 patients, 1048,575 scheduled appointments |
| Viral etiology among children hospitalized for acute respiratory tract infections and its association with meteorological factors and air pollutants: a time-series study (2014-2017) in Macao (Lei et al) | 56 | 2022 | Macao | Neighbourhood heat vulnerability score | Average value per unit, average value of the 5th, 25th, 75th and 95th percentiles, daily mean temps, daily mean humidity, daily mean solar radiation duration, daily mean wind speed, air pollutant data | Macao Meteorological and Geophysical Bureau | Viral etiology for acute respiratory tract infections hospitalisation | ICD codes | Retrospective analysis | 3 | 4880 children aged 1 - 14 years old |
| Vulnerability to episodes of extreme weather: Butajira, Ethiopia, 1998-1999. (Emmelin A. et al) | 57 | 2009 | Ethiopia | Year to year change in weather | Monthly rainfall data from the weather station | National Meteorological Authority, Crop statistics provided by District Agriculture Bureau of the Meskan and Mareko District Authority | Mortality data | Not used | Retrospective analysis | 16 | All deaths registered at the household level |
| Additional outpatient expenditures due to heatwaves: evidence from the Chinese older population (Li et al) | 58 | 2024 | China | Heatwave exposure on outpatient healthcare expenditure | Periods of high temperature above 95th percentile of records, high-resolution grid data set | Local meteorological stations | Outpatient expenditure | Not stated | National survey data, retrospective cohort study | 0 | 22,023 adults over 45 from China 2011-2018” |

***The text in this table is quoted directly from the papers included in this study**
